# Convergent imaging-transcriptomic evidence for disturbed iron homeostasis in Gilles de la Tourette syndrome

**DOI:** 10.1101/2023.05.15.23289978

**Authors:** Ahmad Seif Kanaan, Dongmei Yu, Riccardo Metere, Andreas Schäfer, Torsten Schlumm, Berkin Bilgic, Alfred Anwander, Carol A. Mathews, Jeremiah M. Scharf, Kirsten Müller-Vahl, Harald E. Möller

## Abstract

Gilles de la Tourette syndrome (GTS) is a neuropsychiatric movement disorder with reported abnormalities in various neurotransmitter systems. Considering the integral role of iron in neurotransmitter synthesis and transport, it is hypothesized that iron exhibits a role in GTS pathophysiology. As a surrogate measure of brain iron, quantitative susceptibility mapping (QSM) was performed in 28 patients with GTS and 26 matched controls. Significant susceptibility reductions in the patient cohort, consistent with reduced local iron content, were obtained in subcortical regions known to be implicated in GTS. Regression analysis revealed a significant negative association of tic scores and striatal susceptibility. To interrogate genetic mechanisms that may drive these reductions, spatially specific relationships between susceptibility and gene-expression patterns extracted from the Allen Human Brain Atlas were assessed. Correlations in the striatum were enriched for excitatory, inhibitory, and modulatory neurochemical signaling mechanisms in the motor regions, mitochondrial processes driving ATP production and iron-sulfur cluster biogenesis in the executive subdivision, and phosphorylation-related mechanisms that affect receptor expression and long-term potentiation. This link between susceptibility reductions and normative transcriptional profiles suggests that disruptions in iron regulatory mechanisms are involved in GTS pathophysiology and may lead to pervasive abnormalities in mechanisms regulated by iron-containing enzymes.

## 1. Introduction

Iron is an essential trace element for the vitality of an organism. It is ideally suited for biochemical catalysis due to its ability to transition between two thermodynamically stable oxidation states (Crichton, 2016). This flexibility renders a crucial component of prosthetic groups (e.g., hemes and iron-sulfur clusters) involved in oxygen transport, mitochondrial energy metabolism, cytoskeletal integrity, and neurotransmitter synthesis and transport (Yehuda & Mostofky, 2010). Mediated by the blood-brain barrier, the acquisition of non-heme iron in the brain occurs in an age- and regionally-dependent manner, where its deposition during a “critical period” is necessary for normal brain development (Beard, 2003). This deposition is sharpest in subcortical gray matter (GM), where it overlaps with regions that contain dense proportions of the neurotransmitters dopamine, γ-aminobutyric acid (GABA), and glutamate (Hallgren & Sourander, 1958; Hill, 1988; Li et al., 2014). The atypical homeostasis of iron during different periods of development may affect mechanisms that sustain neurochemical metabolism and myelination (Youdim et al., 1989; Bianco et al., 2010), providing a biological basis for abnormalities in motor and behavioral functions as exhibited by various neuropsychiatric and movement disorders (Beard & Connor, 2003; Stankiewicz et al., 2007; Yehuda, 1990).

Gilles de la Tourette syndrome (GTS) presents an example of a disorder with motor and behavioral deficits as a result of fundamental alterations in the dynamics of cortico-striato-thalamo-cortical circuitry (Felling & Singer, 2011). It is characterized by multiple motor and vocal tics and a high incidence of comorbid features, such as attention deficit/hyperactivity disorder (ADHD), obsessive-compulsive behavior/ disorder (OCB/D), depression, and anxiety (Leckman, 2002). Various genetic studies have indicated that independent allelic variations, each to a small effect, contribute to the manifestation of GTS (Scharf et al., 2013; Yu et al., 2019). Acquired abnormalities in habit formation systems (Delorme et al., 2016; Palminteri et al., 2009; Worbe et al., 2011) are understood to be driven by deficits in subcortical neurochemical signaling (Draper et al., 2014; Fan et al., 2017; Kanaan et al., 2017), thus leading to a burst-like disinhibition of thalamo-cortical output (Mink, 1996; Tremblay et al., 2015). Various work has indicated that patients with GTS exhibit abnormalities in the functional dynamics of tonic and phasic dopaminergic signaling (Grace et al., 2007; Rice et al., 2011; Singer, 2013). These abnormalities are compounded by alterations in the GABAergic (Draper et al., 2014; Kalanithi et al., 2005; Lerner et al., 2012; Tinaz et al., 2014), glutamatergic (Fan et al., 2017; Kanaan et al., 2017), and endocannabinoid (Müller-Vahl et al., 2020) neurotransmitter systems, implying the presence of disturbances in the spatiotemporal dynamics of excitatory, inhibitory, and modulatory subcortical signaling.

One unifying feature exhibited by multiple neurotransmitters is that enzymes involved in their metabolism and the production of their receptors and transporters require iron for typical function. Effects of iron on the dopaminergic system are well recognized as its deficiency leads to deficits in the synthesis, catabolism, transport, and uptake of dopamine (Bianco et al., 2010; Larsen et al., 2020). Other studies have linked iron metabolism to glutamate and GABA, highlighting its role in both excitatory and inhibitory neurotransmitter metabolism and transport (Bianco et al., 2010).

Given these data, it seems plausible that patients with GTS may exhibit an abnormality in the cerebral homeostasis of iron. A link between iron deficiency and an increased risk of contracting neurodevelopmental psychiatric conditions is well established (Chen et al., 2013; Cortese et al., 2008; Degremont et al., 2021; Thomas et al., 2020) as is a link with other movement disorders, such as restless leg syndrome (Connor et al., 2009). This notion of disturbed iron homeostasis is supported by preliminary studies indicating reductions of serum ferritin levels in children (Avrahami et al., 2017; Gorman et al., 2006) and adults (Peterson et al., 1994). Lower ferritin levels were associated with smaller striatal volumes (Gorman et al., 2006), and iron deficiency was suggested to be associated with increased tic severity in children with GTS (Ghosh & Burkman, 2017).

The overarching goal of this work was to investigate the role of iron in GTS pathophysiology and related downstream effects, based on the hypothesis that reduced brain-iron content manifest in adult patients with GTS. We leveraged data available from diverse sources to identify disease-relevant perturbations at multiple scales and provide biological information that is both shared and distinct across modalities. First, we implemented quantitative susceptibility mapping (QSM) as an *in-vivo* proxy measure of brain-iron levels (Deistung et al., 2017; Liu et al., 2015; Möller et al., 2019; Schweser et al., 2011) within well-defined loci of pathophysiology in patients with GTS (*N=*28) and age and gender-matched controls (*N=*26). These data were complemented by a comprehensive clinical assessment battery to identify heterogeneity in symptomatology and by measurements of serum ferritin as a proxy measure of global iron. Additional QSM data from an independent sample of 85 unmatched healthy controls were employed for supplementary analyses. Second, given the conserved and highly stereotyped pattern of cerebral gene expression and its heterogeneity within nuclei subdivisions, we used microarray data from the Allen Human Brain Atlas (AHBA) (Hawrylycz et al., 2012) to assess associations between patterns of gene expression and case-control QSM results. A similar approach has recently been applied to correlate gene expression and cortical iron in Parkinson’s disease (Thomas et al., 2021). The rationale here was that the spatial variation of normative gene expression exhibits a relationship with variations in an image-derived phenotype. As proteins form the backbone of the cellular machinery, we finally explored protein-protein interaction (PPI) networks and the functional enrichment of genes exhibiting maximum covariance with striatal QSM results by uncovering latent variables via partial least squares (PLS) regression.

## 2. Material and methods

### 2.1. Neuroimaging study population

Magnetic resonance imaging (MRI) acquisitions were available from a multi-parametric investigation in 28 adult patients with GTS (18–65 years, 5 female) and 26 adult healthy controls (18–65 years, 8 female), parts of which have been published elsewhere (Gerasch et al., 2016; Kanaan et al., 2017). Imaging data from 23 patients and 26 controls were included in the final analysis following quality control (see below). Both groups were comparable in terms of age, gender and handedness (all right handed; Table 1). Further data from an independent sample of 85 adult healthy controls were used for supplementary investigations (Babayan et al., 2019). All procedures were approved by the Ethics Committees at the Medical Faculty of Leipzig University and Hannover Medical School. All participants gave informed written consent prior to their participation.

**Table 1—.** Demographic and clinical characteristics of the study sample included in the imaging analysis.

| Parameter | Controls | GTS | Test statistics | P-value |
| --- | --- | --- | --- | --- |
| *N* | 26 | 23 |  |  |
| Age in years | 37.92 ± 11.82 | 36.65 ± 10.78 | $t_{47} = 0.38$ | 0.7 |
| Gender (male/female) | 18/8 | 18/5 | OR = 0.52 | 0.36 |
| YGTSS-MS | — | 13.0 ± 4.1 | — | — |
| YGTSS-VS | — | 6.96 ± 5.91 | — | — |
| RVTRS | — | 8.10 ± 4.52 | — | — |
| PUTS | — | 18.9 ± 5.83 | — | — |
| QOL | — | 29.7 ± 20.8 | — | — |
| Y-BOCS | — | 1.34 ± 3.31 | — | — |
| OCI-R | 5.74 ± 5.97 | 14.2 ± 12.5 | $t_{44} = -2.88$ | 0.0062 |
| DSM4 ADHD | 1.17 ± 1.93 | 6.70 ± 5.30 | $t_{44} = -4.63$ | 0.00003 |
| CAARS | 39.27 ± 7.53 | 50.2 ± 12.2 | $t_{44} = -3.49$ | 0.001 |
| MADRS | 0.57 ± 0.77 | 7.96 ± 8.34 | $t_{44} = -4.14$ | 0.0002 |
| BDI-II | 1.78 ± 3.37 | 13.3 ± 12.1 | $t_{44} = -4.27$ | 0.0001 |
| BAI | 2.74 ± 3.29 | 13.3 ± 13.2 | $t_{44} = -3.65$ | 0.0007 |
| Serum ferritin in ng/ml | 196.2 ± 151.3 | 85.6 ± 48.9 | $t_{31} = 2.84$ | 0.0097 |

All recruited subjects were right handed.
**Abbreviations:** ADHD = attention deficit/hyperactivity disorder; BAI = Beck Anxiety Inventory; BDI-II = Beck Depression Inventory II; CAARS = Conners' Adult ADHD Rating Scale; DSM4 = Diagnostic and Statistical Manual of Mental Disorders, Fourth Edition; GTS = Gilles de la Tourette syndrome; MADRS = Montgomery Åsberg Depression Rating Scale; MS = motor tic score; *N* = number of cases; OCI-R = Revised Obsessive-Compulsive Inventory; OR = Fisher exact test odds ratio; *P* = error probability; PUTS = Premonitory Urge for Tics Scale; QOL = quality of life scale; RVTRS = modified Rush Video-Based Tic Rating Scale; *t* = *t*-value; VS = vocal tic score; Y-BOCS = Yale-Brown Obsessive Compulsive Scale; YGTSS = Yale Global Tic Severity Score.

### 2.2. Clinical assessment

All patients underwent a thorough clinical assessment indexing *(i)* tics using the Yale Global Tic Severity Scale (YGTSS), including the total, motor (MS) and vocal tic scores (VS) (Leckman et al., 1989), and the modified Rush Video-Based Tic Rating Scale (RVTRS) (Goetz et al., 1999) as well as premonitory urges using the Premonitory Urge for Tics Scale (PUTS) (Woods et al., 2005); *(ii)* OCB/D using the Yale-Brown Obsessive Compulsive Scale (Y-BOCS) (Goodman et al., 1989) and the Revised Obsessive Compulsive Inventory (OCI-R) (Foa et al., 2002); *(iii)* ADHD using the DSM4 symptom list for ADHD (Saß et al., 2003) and the Conners’ Adult ADHD Rating Scale (CAARS) (Conners et al., 1999); *(iv)* depression using the Montgomery Åsberg Depression Rating Scale (MADRS) (Montgomery & Åsberg, 1977) and the Beck Depression Inventory II (BDI-II) (Beck et al., 1996); and *(v)* anxiety using the Beck Anxiety Inventory (BAI) (Beck et al., 1988). Fourteen were classified into the GTS-only category (without psychiatric comorbidities), whereas the remaining patients exhibited additional OCB/D (*N=*3), ADHD (*N=*5), or the combination of both (*N=*1). Control subjects significantly differed from patients on the scales highlighted in Table 1.

### 2.3. Measurement of serum ferritin

Whenever possible, a 10ml blood sample was collected (*N=*15 patients with GTS and *N*=18 control subjects in the “GTS sample” and *N=*85 subjects in the “independent sample”, respectively) for measuring serum ferritin *in vitro* as a representative measure of the body’s total iron reserves. The sample was first centrifuged at 24,000 rpm for a period of 10 min to separate hematocrit from plasma, which was subsequently stored in 1,000µl aliquots at −70 °C. Serum ferritin levels were quantified based on the electrochemiluminescence immunoassay, in which a voltage applied to a sample containing tagged ferritin molecules induces chemiluminescent emissions that are measured by a photomultiplier (Elecsys 2010, Roche Diagnostics GmbH, Mannheim, Germany).

### 2.4. Image acquisition for QSM

All MRI measurements were performed on a 3T MAGNETOM Verio (Siemens Healthineers, Erlangen, Germany) with a 32-channel head coil. Three-dimensional (3D) *T*_1_-weighted data were acquired using MP2RAGE (Marques et al., 2010) with repetition time, *TR=*5s; echo time, *TE=*3.93ms; inversion times, 0.7 and 2.5s; sagittal slab orientation; matrix 256×256×176; and 1mm isotropic nominal resolution (Streitbürger et al., 2014). Magnetic susceptibility (Δχ) weighted data were acquired with 3D flow-compensated FLASH (flip angle 13°; *TR*=30ms; *TE*=17ms; matrix 256×256×160; 0.8mm isotropic nominal resolution) (Frahm et al., 1986; Haacke & Lenz, 1987).

### 2.5. Image processing for QSM

Details of the image processing pipeline are summarized in Figure 1. An overview of the image processing pipeline is presented in Figure 1. The tools used for reconstructing Δχ maps, Python Magnetic Resonance Tools (PyMRT; https://pypi.org/project/pymrt/) and the Berkeley Advanced Reconstruction Toolbox (BART; https://mrirecon.github.io/bart/), are available as open-source software (Metere & Möller, 2017; Uecker et al., 2021). Briefly, phase maps were reconstructed from multi-channel complex signals using a data-driven coil combination method (Bilgic et al., 2016). It employs singular value decomposition (SVD) to compute a virtual body-coil reference and combines it with an implementation of iTerative Eigenvector-based Self-consistent Parallel Imaging Reconstruction (ESPIRiT) (Uecker et al., 2014). Δχ maps were reconstructed via the superfast dipole inversion approach (Schweser et al., 2013) and were referenced to median cerebrospinal fluid (CSF) susceptibility measured within a subject-specific mask of the lateral ventricles (Straub et al., 2017).

**Figure 1—.**
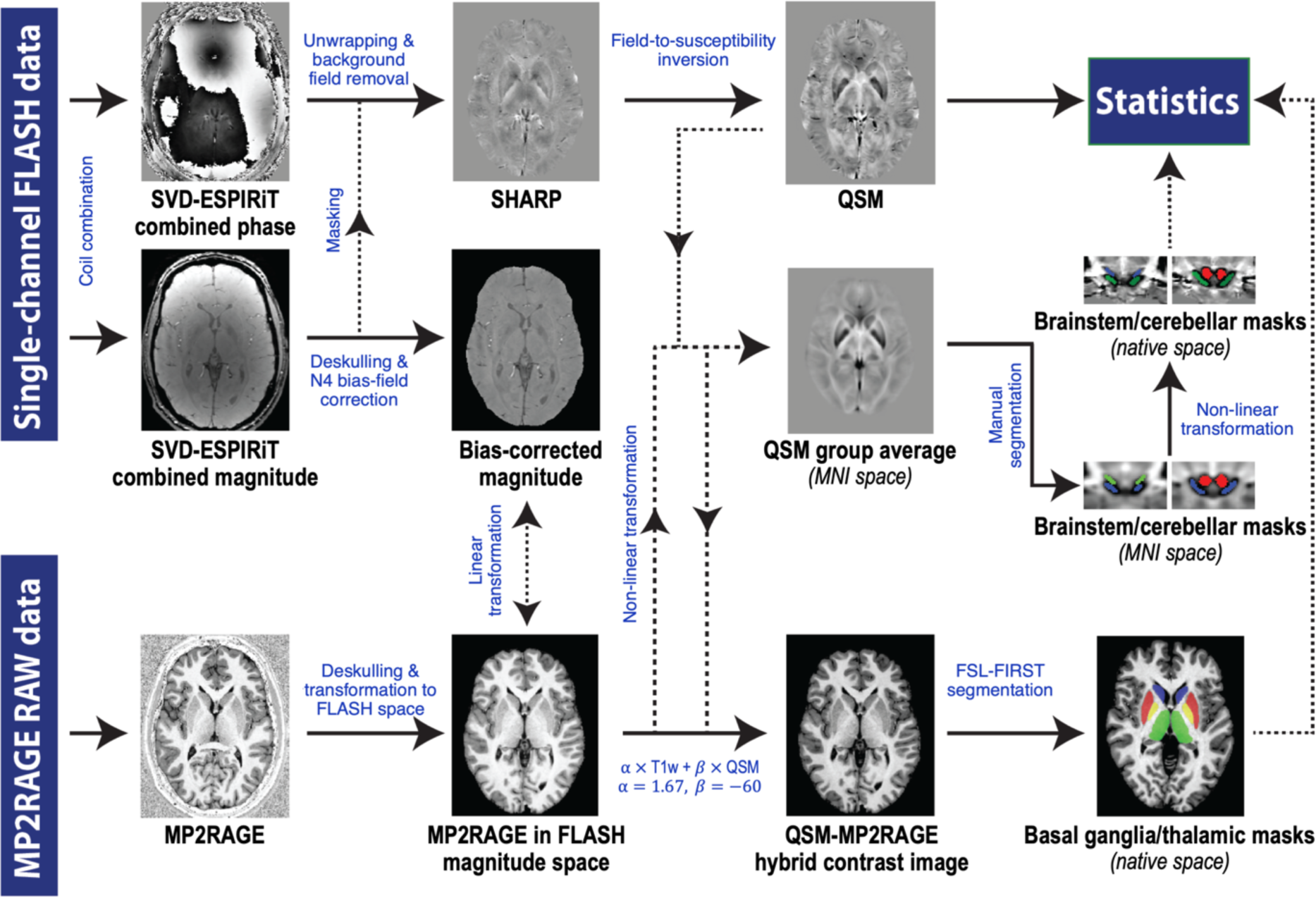
Processing and analysis framework utilized to obtain high-quality quantitative susceptibility maps and subcortical masks. Magnetic susceptibility maps were estimated using the superfast dipole inversion approach, which utilizes Sophisticated Harmonic Artifact Reduction for Phase data (SHARP) to eliminate background-field contributions (Schweser et al., 2013), and thresholded k-space division (TKD) for solving the ill-posed inverse problem from tissue field perturbation to Δχ (Wharton et al., 2010). The sharp deposition of iron-rich structures is clearly visible in basal-ganglia nuclei as well as in brainstem and cerebellar nuclei. The corresponding masks were generated automatically using FSL-FIRST segmentation performed on MP2RAGE-QSM hybrid contrast images (basal ganglia) or by non-linear transformation of atlas-based masks that were carefully delineated on a QSM group-average template in Montreal Neurological Institute (MNI) space (brainstem and cerebellum).

### 2.6. Image quality control

To account for potential differences in the severity of motion-related image artifacts, we used a step-wise, multivariate outlier-detection approach implementing a robust Mahalanobis distance framework (Korkmaz et al., 2014). In particular, low-quality date were removed based on *(i)* image quality indices calculated on the magnitude images and *(ii)* susceptibility values extracted from subcortical nuclei. In general, Mahalanobis distance calculates how far each observation is from the center of a joint distribution, which can be thought of as the centroid in multivariate space. Robust distances are estimated from minimum covariance determinant estimators rather than the sample covariance. Data were regarded as outliers if the robust Mahalanobis distance was greater than the 97.5% quantile of the χ^2^ distribution (Supplementary Figure S1). In the first step, multivariate outliers were detected based on *(i)* the Shannon entropy focus criterion (EFC), which is an index for image ghosting and blurring (Atkinson et al., 1997); *(ii)* the quality index 1 (QI1), which is an index for image degradation resulting from bulk motion, residual magnetization, incomplete spoiling and ghosting (Mortamet et al., 2009); and *(iii)* the smoothness of voxels calculated as the full width at half maximum (FWHM) of the spatial distribution of image intensity values in voxel units (preprocessed-connectomes-project). This step was implemented on the whole sample and identified one severely affected dataset, which was marked for removal (Supplementary Figure S1). To ensure that the remaining datasets did not contain further outliers, multivariate robust squared Mahalanobis distance outlier detection was additionally performed on vectors of median Δχ values extracted from the subcortical masks for each sample separately. This procedure identified four outlier datasets within the patient sample, which were marked for removal (Supplementary Figure S1). Following quality control, group comparisons of magnitude image quality metrics (signal-to-noise ratio; SNR; contrast-to-noise ratio, CNR; foreground-to-background ratio, FBR; EFC; QI1; FWHM) revealed no significant differences between patients and controls (Supplementary Table S1).

### 2.7. Masking of deep GM nuclei

While the QSM data covered the entire brain, the statistical analysis focused on well-defined iron-rich deep-GM and cerebellar nuclei, for which a dominant contribution to magnetic susceptibility from iron is well established (Deistung et al., 2017; Hallgren & Sourander, 1958; Li et al., 2014; Liu et al., 2015). Corresponding masks were generated via *(i)* automated segmentation of hybrid-contrast MP2RAGE-QSM images for the basal ganglia using the FMRIB Software Library (FSL) (Jenkinson et al., 2012) with FMRIB’s Integrated Registration and Segmentation Tool (FIRST) (Patenaude et al., 2011) and *(ii)* non-linear transformation of atlas-based masks for the brainstem (Visser et al., 2016). In particular, masks of the striatum (caudate, putamen), globus pallidus (GP) and thalamus were obtained via the Bayesian model-based subcortical segmentation algorithm implemented in FSL FIRST (Patenaude et al., 2011). It was applied on optimized hybrid-contrast MP2RAGE-QSM images (Deistung et al., 2017; Feng et al., 2017) as outlined in Figure 1 (Kanaan et al., 2018). Robust co-registration between skull-stripped MP2RAGE and FLASH data was achieved using rigid-body linear transformation of the *T*_1_-weighted data onto N4 bias field-corrected FLASH magnitude data employing Advanced Normalization Tools (ANTs; https://github.com/ANTsX/ANTs). Given the difficulty of segmenting brainstem and cerebellar nuclei on *T*_1_-weighted data due to the lack of contrast and the infeasibility of performing manual segmentation of multiple nuclei in many subjects, we utilized an atlas-based registration approach to achieve delineations of brainstem and cerebellar nuclei. Specifically, the diffeomorphic greedy-SyN ANTs non-linear transformation model was used to compute a nonlinear transformation warp between MP2RAGE images and Montreal Neurological Institute (MNI) space. This was used to map each subject’s QSM data into standard space for subsequent calculation of a population-specific average image. The standardized QSM template exhibited high contrast in brainstem and cerebellar regions and was used to carefully delineate masks of the subthalamic nucleus (STN), substantia nigra (SN), red nucleus (RN), and dentate nucleus (DN) (Figure 2). All masks were delineated by the same operator and were subsequently warped back into native QSM space. The same atlas-based registration procedure was applied to obtain subject-specific masks of the lateral ventricles, which were used for referencing the QSM data to CSF (median values). All masks were thresholded at 0.5 to ensure maximal inclusion of GM tissue while limiting partial-volume effects. Following visual inspection of all masks for quality, median susceptibility values from all ROIs were computed for further analysis.

**Figure 2—.**
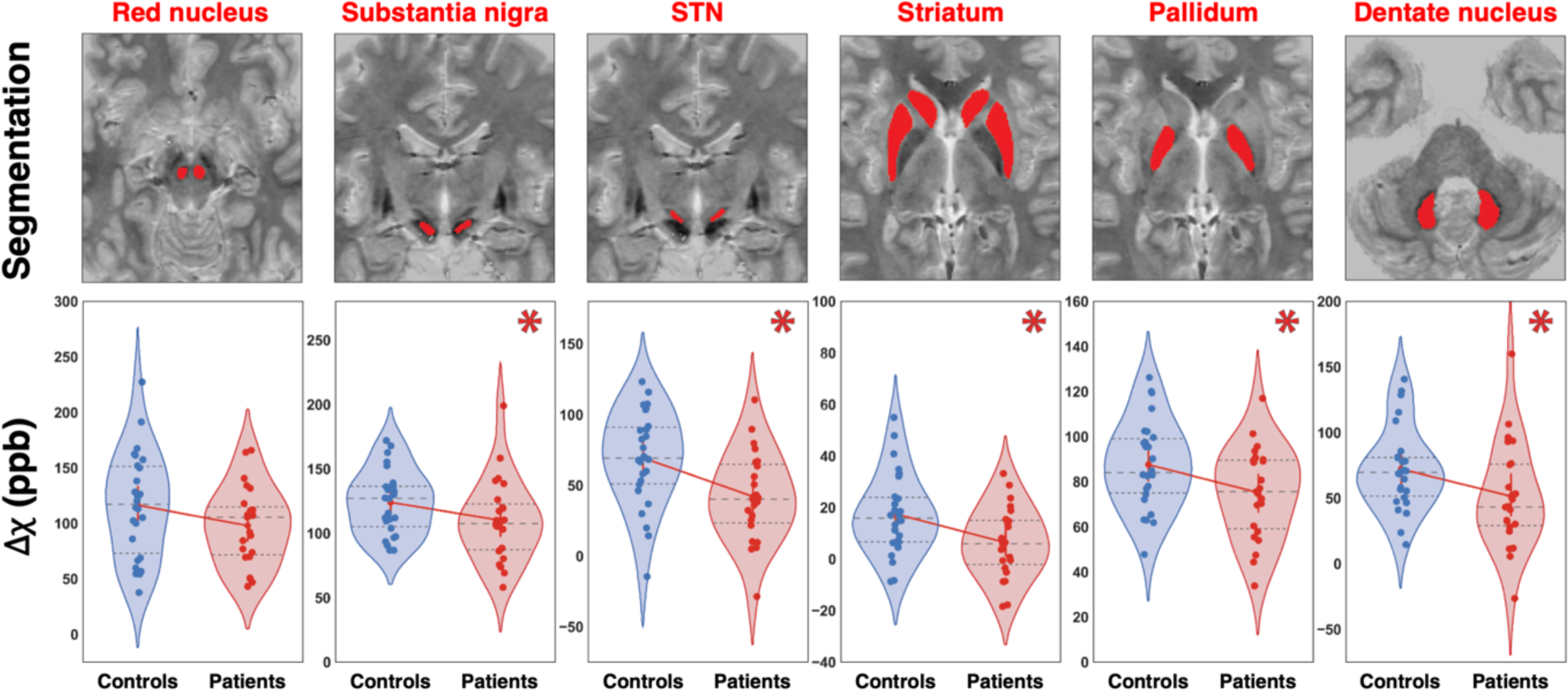
Quantitative susceptibility mapping in GTS. The **upper panel** shows single-subject MP2RAGE-QSM hybrid contrast images illustrating the masking quality of each region of interest (ROI). The **bottom panel** illustrates the probability-density distribution of Δχ values for each ROI. Patients with GTS exhibited significant reductions [false discovery rate (FDR) adjusted *P*-value, *P*FDR<0.05; denoted by *] in the substantia nigra (SN), subthalamic nucleus (STN), striatum, globus pallidus (GP), and dentate nucleus (DN).

### 2.8. Statistical image analysis

Group differences of median Δχ values for each region of interest were assessed using Mann-Whitney-Wilcoxon rank sum tests (significance threshold *P*_FDR_<0.05 after false discovery-rate correction). Group differences in serum ferritin values were assessed using Welch’s *t*-test to account for inhomogeneous variance (Leven’s test). The clinical data were decomposed into a set of low-dimensional scores using principal component analysis (PCA). The correlation matrix revealed sufficient complementarity for data reduction (Figure 3D). Multicollinearity was alleviated by implementing an orthogonal varimax rotation. A multiple linear-regression model accounting for age, gender and image quality indices was used to inspect correlations between iron measures and behavioral principal components (PCs). The variance inflation factor was used to assess multicollinearity between predictor variables.

**Figure 3—.**
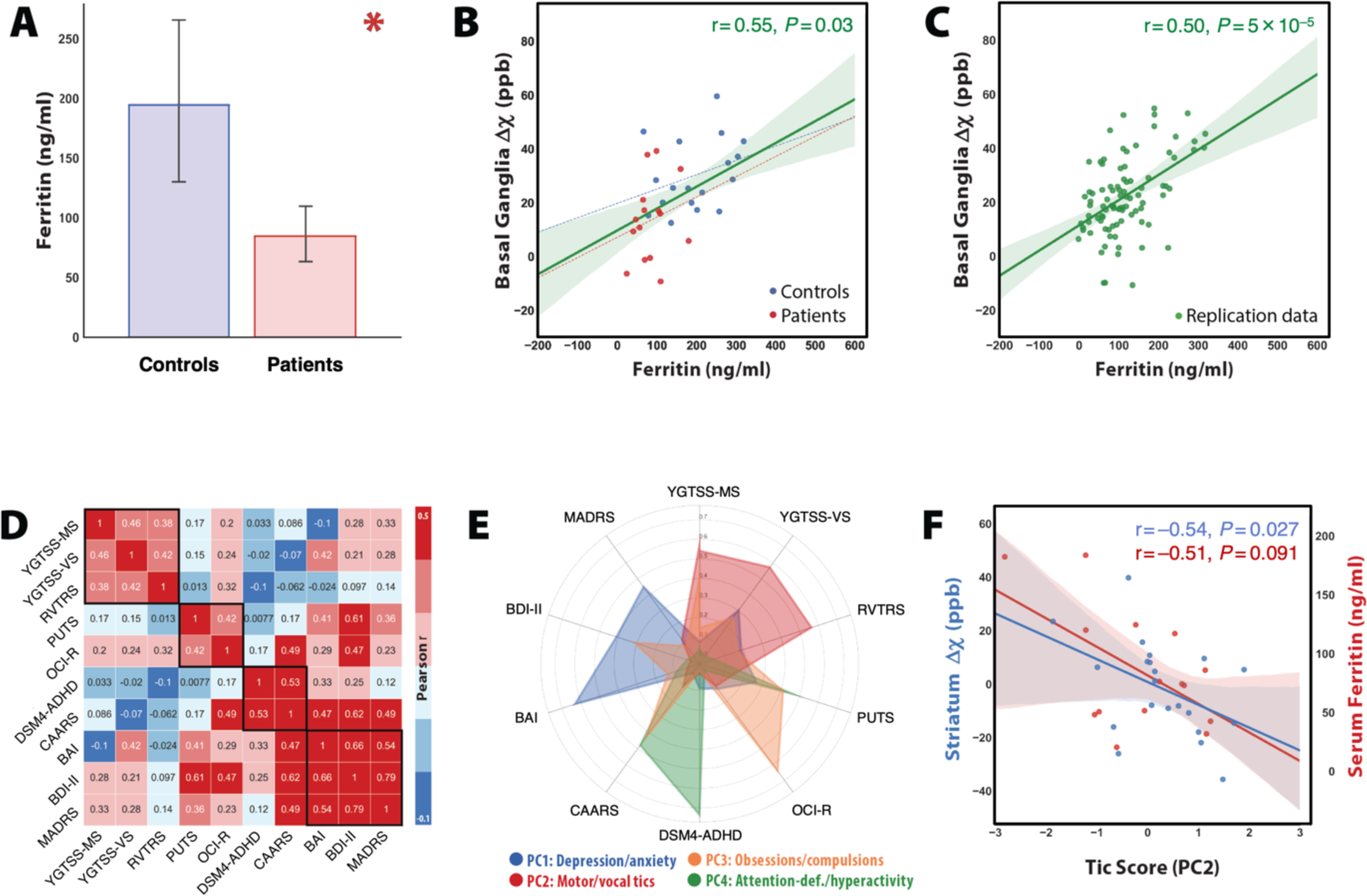
Association between surrogate iron measures and clinical scores. **(A)** Patients with GTS exhibited significant reductions in serum ferritin levels. **(B)** Serum ferritin levels were significantly associated with Δχ of deep GM nuclei in the GTS and control sample. **(C)** This result was replicated in a larger independent sample of healthy volunteers. **(D)** The clinical scores were decomposed using PCA to examine their relationships with iron measures. The correlation matrix between all acquired clinical variables revealed sufficient complementarity for data-reduction. **(E)** PCs with eigenvalues exceeding 1 were extracted yielding a set of four components explaining 77% of the variance. These were interpreted as representative scores of *(i)* depression/anxiety; *(ii)* motor/vocal tics, *(iii)* obsessions/ compulsions, and *(iv)* attention-deficit/hyperactivity as illustrated in the polar plot. **(F)** Regression analysis between PC2 (motor/vocal tics) and iron measures revealed a trend for negative association with serum ferritin levels and a significant negative association with striatal Δχ.

### 2.9. Transcriptional patterning of iron-related striatal variability

The rationale for the analysis of associations between spatial gene expression patterns and Δχ variations as a surrogate of brain iron is illustrated in Figure 4. The default transcriptional architecture is assessed using data from the AHBA (Arnatkevičiūtė et al., 2019; Fornito et al., 2019; Hawrylycz et al., 2012), a high-resolution gene expression atlas obtained from 6 neurotypical individuals (24–57 years, 2 female).

**Figure 4—.**
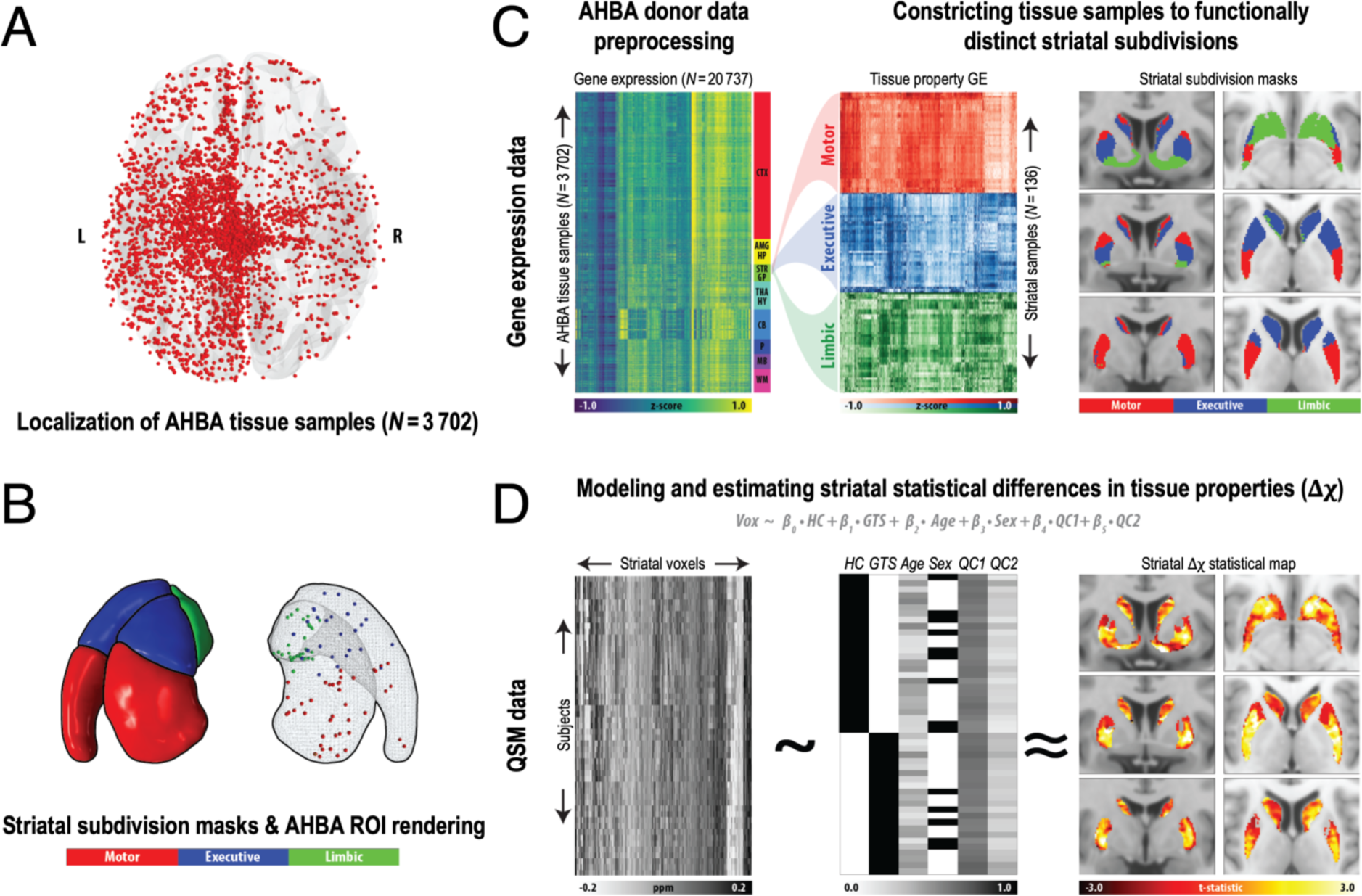
Framework to inspect associations between spatial gene expression profiles and Δχ variations. **(A)** A spatial transcriptomic approach (Arnatkevičiūtė et al., 2019; Fornito et al., 2019) using microarray data from the AHBA was implemented to assess associations between patterns of gene expression and case-control Δχ statistical maps within subdivisions of the striatum as a major locus of pathophysiology in GTS (Kanaan et al., 2017; Lennington et al., 2016; Tremblay et al., 2015). The AHBA contains tissue samples extracted from *N=*3,702 spatially distinct coordinates mapped to MNI space (Morgan et al., 2019; Romero-Garcia et al., 2019; Vértes et al., 2016; Whitaker et al., 2016). **(B)** AHBA tissue samples within the striatum (*N=*136) were defined based on annotation metadata and overlaps between sample coordinates and the subdivisions’ labels of the Harvard-Oxford striatal atlas. **(C)** Gene expression data (*N=*20,737) were then extracted at each coordinate within the striatal motor (*N=*48), executive (*N=*48), and limbic (*N=*40) subdivisions. **(D)** *t*-statistical values of case-control Δχ variations within the same coordinates were modeled using a voxel-wise general linear model that further incorporated age, gender, group, and two indices of image quality. This yielded two matrices representing striatal gene expression (136×20,737) and Δχ *t*-statistical differences (136×1) at 136 positions, which were used to test for enrichment and construct PPI networks following gene weighting via PLS analysis (Morgan et al., 2019; Whitaker et al., 2016).

The combined AHBA and QSM data were used to uncover major genetic classes of variation that exhibit maximum covariance with iron-related differences in GTS. We focused on the striatum, a known major locus of GTS pathophysiology (Felling & Singer, 2011; Gorman et al., 2006; Kanaan et al., 2017), computing voxel-wise Δχ differences within the motor, executive and limbic subdivisions (Figure 4B) and *t*-statistical maps via non-parametric permutation testing with 10,000 permutations using FSL randomize (Figure 4D). Striatal subdivision masks without overlap were derived by transforming the Harvard-Oxford striatal atlas to an average study template before binning the masks to 0.5 followed by one erosion step (Figure 4B). AHBA tissue sample seed masks (1 mm^3^) were created based on standardized AHBA coordinates and mapped to each subdivision, yielding 48 seeds for the motor and executive and 40 seeds for the limbic subdivision (Figure 4C). Ontological AHBA annotations were inspected to ensure that the seeds are mapped within the striatum, and seeds outside of the striatum were removed.

### 2.10. Weighting gene contribution via PLS

To uncover latent associations between Δχ differences and gene-expression patterns, we implemented PLS regression (Morgan et al., 2019; Whitaker et al., 2016). For each striatal functional domain, we constructed matrices exhibiting gene-expression data and case-control Δχ *t*-statistical values from the MRI-defined AHBA tissue coordinates within the motor, executive and limbic subdivisions (Figure 5). Microarray gene-expression matrices (e.g., 48×20,737 for motor division) were used to predict regional Δχ variations as defined by case-control *t*-statistical maps (48×1 for motor division) within each subdivision via PLS regression (Figure 5B,C). In other words, PLS regression was used to identify the linear combination of genes that best predicted the Δχ differences as the response variable. Statistical significance was tested with a two-tailed test at type-I error α=0.05 by permutation of the response variable 1,000 times. Bootstrapping was implemented to assess the error in estimating each gene’s weight in the PLS PCs. The ratio of the weight of each gene to its bootstrap error was used to calculate *Z*-scores and rank the genes according to their contributions to each PLS component (Figure 5).

**Figure 5—.**
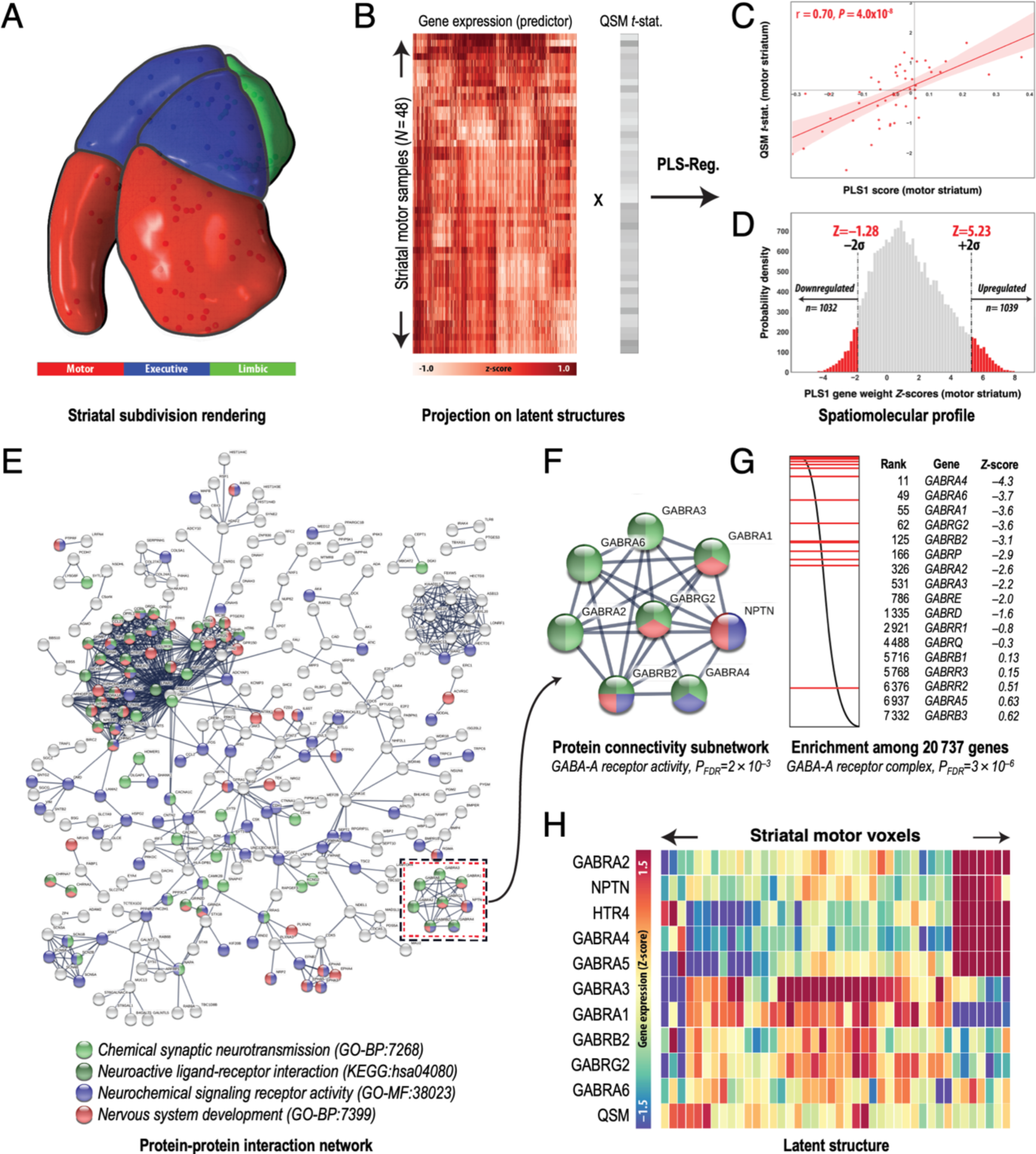
Downregulated genes associated with reduced Δχ and enriched for neurochemical and neurodevelopmental pathways in the motor striatum. **(A)** Transcriptomic and Δχ *t*-statistical data were extracted from colocalized coordinates defined within striatal subdivisions. **(B)** Matrices representing gene expression (48×20,737 predictor variables), and case-control Δχ variation (1×20,737 response variables) were used for cross decomposition via PLS regression. **(C)** Regional scores of the first PLS component (PLS1) exhibited a significant positive correlation with Δχ variations. **(D)** A weighted gene list based on the bootstrapped distribution of PLS1 *Z*-scores was used to define lists of downregulated (PLS^−^) and upregulated (PLS^+^) genes outside the range of the 5% and 95% quantiles of the normal distribution. **(E)** Protein connectivity networks for PLS^−^ genes exhibited significant PPI enrichment (*P*FDR<10^−16^), indicating that they are partially biologically connected. Strongly connected and highly enriched terms included *“chemical synaptic transmission”* (GO-BP:0007268), *“nervous system development”* (GO-BP:0007399), *“signaling receptor activity”* (GO-MF:0038023), and *“neuroactive ligand-receptor interaction”* (KEGG:hsa04080). **(F)** The GO-BP term with the highest PPI network strength (*“GABA signaling pathway”*, GO-BP:0007214) included a sub-network of *“GABA-A receptor activity”* (CL:8612), a result that was replicated when including all 20,737 PLS-weighted genes. **(G)** A supplementary functional enrichment analysis revealed *“GABA-A receptor complex”* (GO-CC:1902711) as the cellular component term with the highest enrichment score. **(H)** The relationship between inhibitory neurotransmission and iron status in GTS is further visible in the heatmap illustrating the underlying structure of GABA receptor expression and case-control Δχ variations.

### 2.11. Gene enrichment and PPI network analysis

Next, we constructed PPI networks to glean information about the network connectivity that contributed jointly toward the underlying biological function. STRING (v11) (Szklarczyk et al., 2019) was used to construct PPI networks for downregulated (denoted as PLS^−^) and upregulated gene sets (PLS^+^) for each component, which were defined as genes outside the 5% and 95% percentiles, respectively (Morgan et al., 2019). Networks were constructed with a minimum interaction score of 0.9 to achieve high confidence of connectivity. ToppFun (Chen et al., 2009) was employed to calculate enrichment of *(i)* gene-ontology (GO) biological processes (BPs), molecular function (MF), and cellular components (CCs) as well as *(ii)* the Kyoto Encyclopedia of Genes and Genomes (KEGG) pathways for downregulated and upregulated genes using a background gene list of 15,745 brain-expressed genes. This list was calculated by excluding probes that did not exceed the background noise in the AHBA dataset by intensity-based filtering (Arnatkevičiūtė et al., 2019; Morgan et al., 2019; Thomas et al., 2021). Multiple-comparison correction of enrichment terms was implemented applying Benjamini and Hochberg’s method for FDR correction. Finally, hierarchical clustering using the Enrichr/Clustergrammer algorithms was employed to visualize the overlap between input genes and GO-biological terms with the highest enrichment score (Kuleshov et al., 2016).

## 3. Results

### 3.1 Group differences of brain-iron measures

Group-level Δχ differences included significant reductions in patients in bilateral brainstem (*U*_47_=184, *P*=0.01), basal ganglia (*U*_47_=167, *P*=0.004) and the combination of all subcortical nuclei (*U*_47_=172, *P*=0.005). These effects were mainly driven by reductions in the striatum (*P*=0.0088), globus pallidus (GP; *P*=0.021), subthalamic nucleus (STN; *P*=0.0026), substantia nigra (SN; *P*=0.032), dentate nucleus (DN; *P*=0.016), and trends for reduction in the red nucleus (*U*_47_=232, *P*=0.091) (Figure 2). Further FDR correction revealed significant Δχ reductions in the striatum, GP, STN, SN, and DN (Cohen’s *d* effect sizes between 0.44 and 0.83 indicating practical significance) (Supplementary Table S2). Patients with GTS showed reduced serum ferritin levels compared to controls (89±49 vs. 196±151 ng/ml; *t*_33_=2.84, *P*=0.0097) (Figure 3A). The serum levels were further correlated with Δχ in the basal ganglia (*r*=0.55, *P*=0.03), which was replicated in the independent sample (*r*=0.5, *P*=5×10^−5^).

### 3.2. Association between brain iron and clinical scores

To inspect correlations between surrogate iron measures and clinical variables, the dimensionality of the clinical data was reduced to four PCs, which—based on the weights of component loadings—were interpreted as representative scores of *(i)* depression/anxiety; *(ii)* motor/vocal tics, *(iii)* OCB/D, and *(iv)* ADHD (Figure 3D,E). Regression analysis for the tic score (PC2) revealed a negative trend with serum ferritin levels (*r* = –0.51, *P*=0.091) and a significant negative association with striatal Δχ (*r* = –0.54, *P*=0.027) (Figure 3F). All correlations exhibited an approximate variance inflation factor of 1.5 indicating little to no multicollinearity between predictor variables.

### 3.3. Transcriptional profiling of Δχ reductions

PLS regression was implemented to identify the patterns of gene expression that most strongly correlated with the distribution of striatal Δχ reductions. The first component (PLS1) explained 48.2%, 52.2%, and 49.1% of the variance in the case-control Δχ *t*-statistical maps of the motor, executive, and limbic subdivisions, respectively, which was significantly more than expected by chance (permutation test, *P<*0.001). For all subdivisions, PLS1 gene expression scores exhibited strong positive correlations with case-control Δχ variations (*r*≈0.7, *P<*10^−6^) indicating a spatial relationship between normative gene expression and Δχ in GTS (Figure 5C, Supplementary Figure S2 and Supplementary Table S3). Hence, more positively weighted genes exhibited overexpression and more negatively weighted genes exhibited underexpression in striatal regions, in which the patients had increased and decreased Δχ, respectively. Given the strong correlation and variance level explained by this component, we only considered PLS1 gene-expression weights for further analysis.

Multiple univariate tests and bootstrapping were used to evaluate whether the weights of genes on PLS1 were significantly different from zero. Genes with *Z*-scores (*P*_FDR_<0.05) outside the range of the 5^th^ and 95^th^ percentiles of the weighted gene list were regarded as exhibiting underexpression and overexpression, respectively, as a result of case-control Δχ variations. For the motor striatum, the null hypothesis was refuted for 1,032 PLS^−^ genes (*Z* < –1.28) and 1,039 PLS^+^ genes (*Z* > 5.23) (Figure 5D and Supplementary Figure S2). A correspondence was exhibited for gene counts of the executive and limbic subdivisions (Supplementary Figures S3 and S4).

Each set of PLS^−^ and PLS^+^ genes of each striatal subdivision was used to construct PPI networks using STRING (Figure 5E). Significant PPI enrichment was observed for PLS^−^ genes of the motor striatum and for PLS^+^ genes for the motor, executive and limbic subdivisions (Supplementary Table S4). The PPI networks exhibited high enrichment scores (*P<*10^−8^) and moderate local clustering coefficients (≈0.3). This indicates that the spatiomolecular profile of PLS genes associated with Δχ variation exhibited more interactions among themselves than would be expected by a random set of genes and, therefore, partial biological connectivity.

PLS^−^ genes associated with case-control Δχ variation in the motor striatum demonstrated enrichment for BPs and pathways involved in excitatory, inhibitory and modulatory neurochemical systems (Supplementary Figure S5). Non-overlapping enrichment terms included *“chemical synaptic transmission”* (GO-BP:0007268), *“GABAergic synaptic transmission”* (GO-BP:0051932), *“positive regulation of actin cytoskeleton reorganization”* (GO-BP:2000251), and *“neuroactive ligand-receptor interaction”* (KEGG:hsa04080). Hierarchical clustering of PLS^−^ genes among GO-BP terms demonstrated an overlap for genes encoding subunits of GABA receptor complexes, ionotropic *N*-methyl-D-aspartate (NMDA) glutamatergic receptors, and the dopaminergic receptor D2 (Figure 6A). On a cellular level, we observed significant enrichment of *“integral”* and *“intrinsic”* pre- and postsynaptic membrane components (Supplementary Figure S6) indicating the involvement of receptor complex proteins. Considering PLS^+^ genes in the motor striatum, we observed enrichment of fairly broad translational and metabolic process terms that were not analyzed further.

**Figure 6—.**
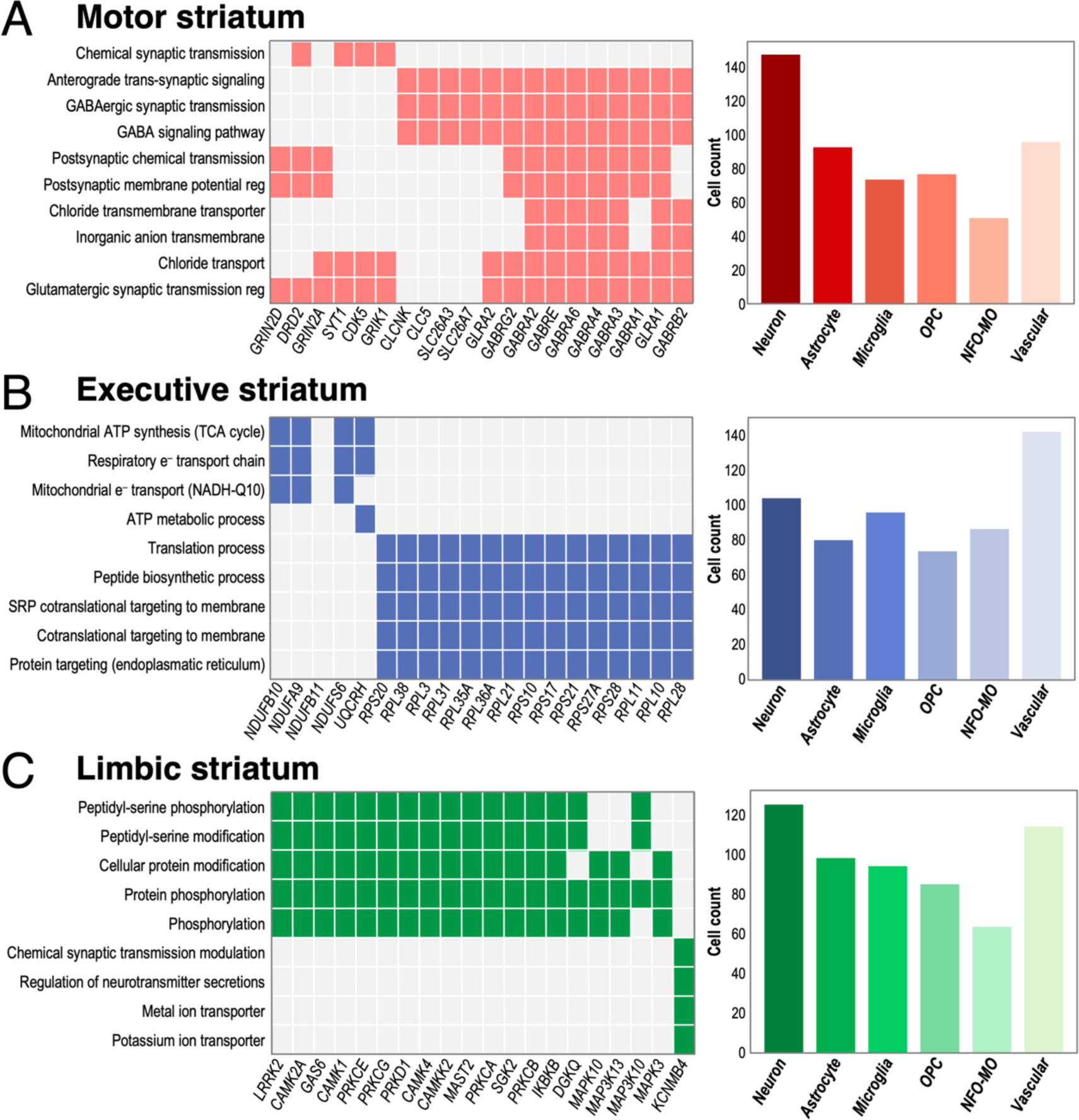
Gene overlap among the highest enriched terms and the differential expression of marker genes across cell types. **(A)** Hierarchical-clustering results obtained with PLS^−^ in the motor striatum, demonstrating an overlap for genes encoding subunits of GABA receptor complexes (e.g., GABRA1, GABRB), glutamate NMDA receptor (e.g., GRIN2A, GRIN2D) or dopamine D2 receptor (e.g., DRD2). **(B)** Results for PLS^+^ in the executive striatum, with overlap of genes encoding inner-mitochondrial membrane subunits of NADH dehydrogenase and cytochrome B (e.g., NDUFA9, UQCRH) or ribosomal complex subunits (e.g., RPS27A, RPL38). **(C)** Results for PLS^+^ in the limbic striatum highlighting an involvement of subterms for *“iron incorporation into iron-sulfur cluster”* processes with clustered genes including LRRK2, implicated in Parkinson’s disease, or calcium-calmodulin dependent CAMKK2, triggering the activity of varied kinases and regulating transferrin trafficking and turnover (Sabbir, 2018, 2020). The cellular affiliation score to each gene in the gene lists that is shown in the right column was assigned according to prior criteria from Zhang *et al*. (2014). Abbreviations: NFO-MO = formed and myelinating oligodendrocytes, OPC: oligodendrocyte progenitor cells.

For the executive striatum, PLS^+^ genes exhibited enrichment for two BP clusters involved in mitochondrial gene expression and energy production as well as for mitochondrial CC terms (Supplementary Figures S7 and S8). These results were particularly exemplified by genes encoding the iron-sulfur replete inner-mitochondrial membrane subunits of NADH dehydrogenase and cytochrome B and ribosomal complex subunits (Figure 6B).

In the limbic striatum, PLS^+^ genes highlighted regulatory processes involved in intracellular signal transduction, neurotransmitter secretion and synaptic plasticity (Supplementary Figures S9 and S10). Top enriched KEGG pathways included *“long-term potentiation”* (KEGG:hsa04720). Relatedly, phosphorylation related GO-MF terms included *“calmodulin binding”* (GO-MF:0005516) and *“calmodulin-dependent protein kinase activity”* (GO-MF:0004683). Hierarchical clustering of genetic overlap between upregulated PLS^+^ gene and the top enriched GO-BP terms included kinases also involved in other movement disorders and the trafficking and turnover of transferrin (Figure 6C).

## 4. Discussion

To investigate a role of iron in GTS pathophysiology, we combined QSM targeted at subcortical iron content and a spatiomolecular approach to interrogate GO enrichment relations with striatal Δχ variations. Whereas QSM is sensitive to both para- and diamagnetic materials, most notably iron and myelin, respectively, the iron contribution dominates that of myelin in subcortical structures (Langkammer et al., 2012; Möller et al., 2019). Most brain iron is stored in ferritin containing a core of up to 4,500 Fe^3+^ ions. Its superparamagnetic susceptibility and, thus, effective magnetic field perturbation can be readily detected with QSM *in vivo* (Möller et al., 2019), allowing to go beyond previous reports of reduced (global) serum ferritin levels in patients with GTS (Figure 3A). Consistently, our data demonstrate Δχ reductions in the SN, STN, striatum, GP, and DN (Figures 2 and 3B) as well as an association of striatal Δχ with tic severity (Figure 3F). This suggests that patients with GTS exhibit disturbances in iron homeostasis in subcortical regions that typically exhibit the sharpest concentrations of iron (Hallgren & Sourander, 1958; Li et al., 2014) and, at the same time, the densest proportions of dopamine, GABA, and glutamate. Spatial transcriptional patterns of gene expression yielded correlations with striatal Δχ variations and were enriched for *(i)* excitatory, inhibitory, and modulatory neurochemical signaling mechanisms in the motor regions, *(ii)* mitochondrial processes driving ATP production and iron-sulfur cluster biogenesis in the executive subdivision, and *(iii)* phosphorylation-related mechanisms that affect receptor expression and long-term potentiation. As the developmental processes of synaptogenesis, dendritogenesis and myelination are highly dependent on iron-containing enzymes, subtle deficiencies of iron content throughout the lifespan may ultimately influence mechanisms of subcortical neurochemical signaling and drive the acquisition and persistence of deficits in motor behavior.

### 4.1. Disturbed iron homeostasis

Investigations of the developing brain indicate that iron deficiency may lead to a specific set of behavioral outcomes that are dependent on its timing, severity, and duration (Beard, 2003; Lozoff & Georgieff, 2006). Human infant and animal studies have consistently shown that early life iron deficiency leads to long-term abnormalities in motor, cognitive, and affective behavior that are irreversible with iron repletion at weaning (Beard & Connor, 2003; Kennedy et al., 2016; Lozoff et al., 2006). Children and adults with perinatal, neonatal, and postnatal iron deficiency exhibited cognitive impairments in selective attention, perceptual speed, and inhibitory control, in addition to depressive and anxiety-like symptoms (Kennedy et al., 2016). Within the motor domain, early-life iron deficiency was associated with delays in developmental motor milestones and lower global indices of motor function (Kennedy et al., 2016).

Some of these symptoms also manifest in GTS, including abnormalities in syntactic motor chains, inhibitory control, selective attention, and depressive/anxiety-like behavior (Leckman, 2002; Lozoff & Georgieff, 2006). The correspondence in exhibited behavior between iron deficiency and GTS might be reflected by similarities in their etiological basis. Considering that GTS is, in essence, a polygenic disorder with environmental influences (Deng et al., 2012; Hoekstra et al., 2013), variants in genes involved in the regulation of iron metabolism and environmental factors influencing iron availability have both been reported. In a microarray study exploring the influence of gestational/lactational iron deficiency on mRNA expression levels, variants in genetic clusters involved in cytoskeletal stability and synaptic function exhibited chronic downregulations following iron repletion therapy at weaning (Clardy et al., 2006). Similarly, two independent studies demonstrated associations between GTS and variants in the BTDB9 gene, which contains a PPI motif (BTB domain) implicated in cytoskeletal regulation, ion channel gating, and transcriptome repression (Guo et al., 2012; Rivière et al., 2009). The cytoskeleton forms the primary backbone of neuronal architecture, in which the coordinated interplay of cytoskeletal elements is essential for axonal outgrowth, synapse formation and cargo logistics (Goellner & Aberle, 2012). Cytoskeletal abnormalities driven by disturbances in iron homeostasis may, hence, affect synaptic efficacy and underlie some of the long-term outcomes in GTS. On the other hand, gestational noxious exposures including maternal smoking and prenatal stressors have been linked to increased risk of both iron deficiency and the onset of tics (Deng et al., 2012; Hoekstra et al., 2013). This indicates that deficits in iron homeostasis might impact the onset and course of GTS and might be driven by a complex relationship between genetic and environmental influences.

Previous preliminary work suggested an association of iron deficiency and increased tic severity in children with GTS as well as a trend towards improved tic severity scores upon follow-up on iron supplementation therapy (Ghosh & Burkman, 2017). For a definite conclusion about iron therapy, randomized placebo-controlled prospective studies are required. Such trials may benefit from integrating QSM as a potential biomarker assessing local iron content—and, hence, therapy response—in brain regions assumed to be involved in tic generation.

Although early-life iron deficiency is associated with long-term deficits in motor and non-motor behavior, this does not necessarily last into adulthood. In longitudinal studies assessing early-life iron deficiency over 19 years, deficiency in infancy led to persistent motor and non-motor deficits, despite being present in less than 5% of the subjects after 19 years (Lozoff & Georgieff, 2006; Lukowski et al., 2010). However, a subtle iron-deficient status seems to persist throughout the lifespan in GTS considering reduced serum ferritin in pediatric samples reported previously (Avrahami et al., 2017; Gorman et al., 2006) as well as reductions of subcortical Δχ observed in the current work. Remarkably, previous histological studies (Hallgren & Sourander, 1958) as well as QSM *in vivo* (Li et al., 2014; Persson et al., 2015) observed that brain iron content in many deep GM nuclei (i.e., regions showing significant Δχ reductions in our cohort of adult patients with GTS) showed rapid growth from birth until about 20 years of age, approaching a plateau at middle age. Although no direct evidence can be derived from these trajectories to interpret our observations, it seems plausible that decreased Δχ in adult patients reflects persistent iron deficiency during development. While early-life iron deficiency may lead to severe abnormalities in neurochemical mechanisms involved in developmental processes (e.g., habit formation), adult iron deficiency may exacerbate abnormalities in neurochemistry.

The associations between Δχ reductions and gene expression patterns suggest that disruptions in mechanisms underpinning homeostatic iron levels in GTS are associated with abnormalities in mechanisms sustaining the typical spatiotemporal dynamics of striatal neurochemical signaling. Dopamine exerts a powerful influence over striatal output by modulating cortico-striatal afferents innervating distinct populations of striato-nigral and striato-pallidal medium spiny neurons. The spatiotemporal synergy between tonic and phasic dopaminergic signals emanating from the SN and ventral tegmental area is a key factor in driving thalamo-cortical output and coordinated behavior, probably due to the integrated output of D1- and D2-receptor medium spiny neurons (Grace et al., 2007). As such, alterations in the mechanisms sustaining this typical spatiotemporal synergy may have a profound influence on reinforcement learning and habit formation systems that are governed by striatal neurons coding the serial order of syntactic natural behavior (Aldridge et al., 1998; Cromwell & Berridge, 1996).

Among disruptions in elements sustaining typical tonic/phasic dopaminergic signaling, alterations in the D1/D2 receptor density, dopamine transporter function, and monoamine catabolism have been reported in GTS (Singer, 2013) and also observed in association with iron deficiency (Lozoff, 2011). This implies a role of iron deficiency in abnormal dopaminergic neurotransmission, which is supported by observed Δχ reductions in the SN (Lozoff, 2011). However, alterations in tonic/phasic dopaminergic signaling may also be driven by deficits in glutamatergic and GABAergic afferent systems regulating SN dopaminergic output towards the striatum (Grace et al., 2007). Recently reported alterations in striatal glutamate levels (Kanaan et al., 2017) and the association of reduced striatal Δχ and highly enriched terms related to GABA signaling observed here indicate that iron might influence GABA-glutamate-glutamine cycling. Consistently, previous work indicated that iron deficiency during gestational and lactational periods in rodents leads to alterations in key enzymes involved in the GABA-glutamate-glutamine cycle, ultimately leading to reductions in striatal, pallidal, and hippocampal GABAergic/glutamatergic neurotransmission (Anderson et al., 2007; Erikson et al., 2002; Ward et al., 2007).

### 4.2. Limitations and future directions

Besides iron sequestered in ferritin, which is considered to be the dominant contribution to QSM in most subcortical brain regions, neuromelanin efficiently binds iron and is abundant in dopaminergic neurons of the SN and locus coeruleus (Möller et al., 2019). Further contributions may result from copper(II), manganese, or zinc (Wallstein et al., 2022). Abnormalities in the homeostasis of these ions might contribute to GTS pathophysiology––given their roles in neurochemical signaling. However, we do not expect major bias on Δχ estimates given their relative scarcity compared to iron (Krebs et al., 2014). Brain transferrin levels are one order of magnitude below those of ferritin and probably too low to be detected by QSM (Möller et al., 2019). Hence, alterations (e.g., upregulation) of transferrin levels cannot be assessed with our data.

While previous work established relationships between iron and GABA/dopamine metabolism, we obtained only indirect links without direct measurements. Similarly, the AHBA is limited to normative transcriptomics profiling. Patient-directed atlases, if available in future work, could be extremely useful for provide direct links to GTS transcriptomics.

Only few patients in our cohort were classified as exhibiting additional ADHD or OCB/D or both. These numbers are too small to determine how iron metrics could be associated with corresponding non-motor profiles or whether there are distinct spatiomolecular profiles of GTS and its comorbidities. Brain iron homeostasis may also correlate in other neurological disorders, specifically those having dopaminergic system involvement, and not exclusively GTS. Therefore, future work is needed to address the specificity of patterns of regional brain iron disruption as well their role in network dysfunction considering the current notion of characterizing neuropsychiatric disorders (or GTS and tic disorders in particular) as “network disorders” rather than resulting from damage to isolated brain regions (Fox, 2018; Ganos et al., 2022).

Another limitation regarding our study population is that our conclusions are based on a cohort of adult age, whereas GTS is a childhood neuropsychiatric disorder. Studies of possible changes in brain iron homeostasis in children with GTS are, therefore, needed to test the assumption that early-life iron deficiency may contribute to manifestation of GTS. Ideally, a longitudinal follow-up could help to further disentangle the relationship between tic severity and brain iron levels across the lifespan. While such studies are difficult to perform, QSM—or MRI in general—is an ideal method for targeting such aspects due to its completely noninvasive nature and the ability to provide quantitative data. More generally, we advocate replication of our study in an independent sample. In addition to using QSM as a proxy for iron homeostasis, such work might be combined with proton spectroscopy of glutamate and GABA or positron emission tomography of dopamine signaling for a multimodal approach to characterize GTS pathophysiology.

## 5. Conclusions

Disruptions in iron regulatory mechanisms––presumably driven by relations between genetic and environmental influences––may contribute to the establishment of GTS and lead to pervasive abnormalities in mechanisms regulated by iron-containing enzymes. Abnormalities in mRNA transcription, cytoskeletal regulation, synapse formation, axonal outgrowth, ion-channel gating, iron-sulfur cluster biogenesis mechanisms may lead to alterations in the spatiotemporal dynamics of excitatory, inhibitory, and modulatory subcortical signaling, thus driving the manifesting clinical features in GTS.

## Data Availability

Gene weights and enrichment data produced in the present study are available upon reasonable request to the authors.

## Acknowledgements

This work was funded by the European Commission through the Marie Skłodowska-Curie ITNs “TS-EUROTRAIN” (FP7-PEOPLE-2012-ITN, Grant No. 316978) to KMV and “HiMR” (FP7-PEOPLE-2012-ITN, Grant No. 316716) and “INSPiRE-MED” (H2020-MSCA-ITN-2018, Grant No. 813120) to HEM and, in part, by the Helmholtz Alliance “ICEMED—Imaging and Curing Environmental Metabolic Diseases” (HA-314) to HEM. BB acknowledges funding by the National Institutes of Health (Grant Nos. R01 EB028797, R03 EB031175, U01 EB026996, U01 EB025162, and P41 EB030006). CAM and JMS acknowledge funding by the National Institutes of Health (Grant Nos. R01 NS102371 and R01 NS105746). Preliminary results of this work were presented at the 26^th^ Annual Meeting of the International Society for Magnetic Resonance in Medicine (ISMRM) in Paris, France. The authors thank Sarah Gerasch for performing the clinical assessments, Karla Claudio for helpful discussions, the German Tourette Society advocacy groups TGD e. V. and IVTS e. V. for their support in patient recruitment, and all patients and control subjects for their participation.

## References

Aldridge, J.W., Berridge, K.C., 1998. Coding of serial order by neostriatal neurons: A ‘natural action’ approach to movement sequence. J. Neurosci. 18, 2777–2787. https://doi.org/10.1523/JNEUROSCI.18-07-02777.1998.

Anderson, J.G., Cooney, P.T., Erikson, K.M., 2007. Brain manganese accumulation is inversely related to γ-amino butyric acid uptake in male and female rats. Toxicol. Sci. 95, 188–195. https://doi.org/10.1093/toxsci/kfl130.

Arnatkevičiūtė, A., Fulcher, B.D., Fornito, A., 2019. A practical guide to linking brain-wide gene expression and neuroimaging data. NeuroImage 189: 353–367. https://doi.org/10.1016/j.neuroimage.2019.01.011.

Atkinson, D., Hill, D.L., Stoyle, P.N., Summers, P.E., Keevil, S.F., 1997. Automatic correction of motion artifacts in magnetic resonance images using an entropy focus criterion. IEEE Trans. Med. Imaging 16, 903–910. https://doi.org/10.1109/42.650886.

Avrahami, M., Barzilay, R., HarGil, M., Weizman, A., Watemberg, N., 2017. Serum ferritin levels are lower in children with tic disorders compared with children without tics: A cross-sectional study. J. Child Adolesc. Psychopharmacol. 27, 192–195. https://doi.org/10.1089/cap.2016.0069.

Babayan, A., Erbey, M., Kumral, D., Reinelt, J.D., Reiter, A.M.F., Röbbig, J., Schaare, H.L., Uhlig, M., Anwander, A., Bazin, P.-L., Horstmann, A., Lampe, L., Nikulin, V.V., Okon-Singer, H., Preusser, S., Pampel, A., Rohr, C.S., Sacher, J., Thöne-Otto, A., Trapp, S., Nierhaus, T., Altmann, D., Arelin, K., Blöchl, M., Bongartz, E., Breig, P., Cesnaite, E., Chen, S., Cozatl, R., Czerwonatis, S., Dambrauskaite, G., Dreyer, M., Enders, J., Engelhardt, M., Fischer, M.M., Forschack, N., Golchert, J., Golz, L., Guran, C.A., Hedrich, S., Hentschel, N., Hoffmann, D.I., Huntenburg, J.M., Jost, R., Kosatschek, A., Kunzendorf, S., Lammers, H., Lauckner, M.E., Mahjoory, K., Kanaan, A.S., Mendes, N., Menger, R., Morino, E., Näthe, K., Neubauer, J., Noyan, H., Oligschläger, S., Panczyszyn-Trzewik, P., Poehlchen, D., Putzke, N., Roski, S., Schaller, M.-C., Schieferbein, A., Schlaak, B., Schmidt, R., Gorgolewski, K.J., Schmidt, H.M., Schrimpf, A., Stasch, S., Voss, M., Wiedemann, A., Margulies, D.S., Gaebler, M., Villringer, A., 2019. A mind-brain-body dataset of MRI, EEG, cognition, emotion, and peripheral physiology in young and old adults. Sci. Data 6, 180308. https://doi.org/10.1038/sdata.2018.308.

Beard, J., 2003. Iron deficiency alters brain development and functioning. J. Nutr. 133, 1468S–1472S. https://doi.org/10.1093/jn/133.5.1468S.

Beard, J.L., Connor, J.R., 2003. Iron status and neural functioning. Annu. Rev. Nutr. 23, 41–58. https://doi.org/10.1146/annurev.nutr.23.020102.075739.

Beck, A.T., Epstein, N., Brown, G., Steer, R.A., 1988. An inventory for measuring clinical anxiety: Psychometric properties. J. Consult. Clin. Psychol. 56, 893–897. https://doi.org/10.1037/0022-006X.56.6.893.

Beck, A.T., Steer, R.A., Brown, G.K., 1996. Manual for the Beck Depression Inventory-II. Psychological Corporation, San Antonio, TX.

Bianco, L., Unger, E., Beard, J., 2010. Iron deficiency and neuropharmacology, in: Yehuda, S., Mostofsky, D. (Eds.), Iron Deficiency and Overload. Humana Press, Ney York, NY, pp. 141–158. https://doi.org/10.1007/978-1-59745-462-9_8.

Bilgic, B., Polimeni, J.R., Wald, L.L., Setsompop, K., 2016. Automated tissue phase and QSM estimation from multichannel data. Proceedings of the 24th Annual Meeting of ISMRM, Singapore, abstract 2849.

Chen, J., Aronow, B.J., Jegga, A.G., 2009. Disease candidate gene identification and prioritization using protein interaction networks. BMC Bioinformatics 10, 73. https://doi.org/10.1186/1471-2105-10-73.

Chen, M.-H., Su, S.-T., Chen, Y.-S., Hsu, Y.-W., Huang, K.-L., Chang, W.-H., Chen, T.-J., Bai, Y.-M., 2013. Association between psychiatric disorders and iron deficiency anemia among children and adolescents: A nationwide population-based study. BMC Psychiatry 13, 161. https://doi.org/10.1186/1471-244X-13-161.

Clardy, S.L., Wang, X., Zhao, W., Liu, W., Chase, G.A., Beard, J.L., Felt, B.T., Connor, J.R., 2006. Acute and chronic effects of developmental iron deficiency on mRNA expression patterns in the brain, in: Parvez, H., Riederer, P. (Eds.). Oxidative Stress and Neuroprotection. J. Neural. Transm. Suppl. 71, 173–196. https://doi.org/10.1007/978-3-211-33328-0_19.

Connor, J.R., Wang, X.-S., Allen, R.P., Beard, J.L., Wiesinger, J.A., Felt, B.T., Earley, C.J., 2009. Altered dopaminergic profile in the putamen and substantia nigra in restless leg syndrome. Brain 132, 2403–2412. https://doi.org/10.1093/brain/awp125.

Conners, C.K., Erhardt, D., Sparrow, E.P., 1999. Conner’s Adult ADHD Rating Scales: Technical manual. Multi-Health Systems, Inc. (MHS), North Tonawanda, NY.

Cortese, S., Lecendreux, M., Dalla Bernardina, B., Mouren, M.C., Sbarbati, A., Konofal, K., 2008. Attention-deficit/hyperactivity disorder, Tourette’s syndrome, and restless legs syndrome: The iron hypothesis. Med. Hypotheses 70, 1128–1132. https://doi.org/10.1016/j.mehy.2007.10.013.

Crichton, R., 2016. Iron Metabolism: From Molecular Mechanisms to Clinical Consequences, 4th edition. Wiley, Chichester, UK. https://doi.org/10.1002/9781118925645.

Cromwell, H.C., Berridge, K.C., 1996. Implementation of action sequences by a neostriatal site: A lesion mapping study of grooming syntax. J. Neurosci. 16, 3444–3458. https://doi.org/10.1523/JNEUROSCI.16-10-03444.1996

Degremont, A., Jain, R., Philippou, E., Latunde-Dada, G.O., 2021. Brain iron concentrations in the pathophysiology of children with attention deficit/hyperactivity disorder: A systematic review. Nutr. Rev. 79, 615–626. https://doi.org/10.1093/nutrit/nuaa065.

Deistung, A., Schweser, F., Reichenbach, J.R., 2017. Overview of quantitative susceptibility mapping. NMR Biomed. 30, e3569. https://doi.org/10.1002/nbm.3569.

Deng, H., Gao, K., Jankovic, J., 2012. The genetics of Tourette syndrome. Nat. Rev. Neurol. 8, 203–213. https://doi.org/10.1038/nrneurol.2012.26.

Delorme, C., Salvador, A., Valabrègue, R., Roze, E., Palminteri, S., Vidailhet, M., de Wit., S., Robbins, T., Hartmann, A., Worbe, Y., 2016. Enhanced habit formation in Gilles de la Tourette syndrome. Brain 139, 605–615. https://doi.org/10.1093/brain/awv307.

Draper, A., Stephenson, M.C., Jackson, G.M., Pépés, S., Morgan, P.S., Morris, P.G., Jackson, S.R., 2014. Increased GABA contributes to enhanced control over motor excitability in Tourette syndrome. Curr. Biol. 24, 2343–2347. https://doi.org/10.1016/j.cub.2014.08.038.

Erikson, K.M., Shihabi, Z.K., Aschner, J.L., Aschner, M., 2002. Manganese accumulates in iron-deficient rat brain regions in a heterogeneous fashion and is associated with neurochemical alterations. Biol. Trace Elem. Res. 87, 143–156. https://doi.org/10.1385/BTER:87:1-3:143.

Fan, S., Cath, D.C., van den Heuvel, O.A., van der Werf, Y.D., Schöls, C., Veltman, D.J., Pouwels, P.J.W., 2017. Abnormalities in metabolite concentrations in Tourette’s disorder and obsessive-compulsive disorder—A proton magnetic resonance spectroscopy study. Psychoneuroendocrinology 77: 211–217. https://doi.org/10.1016/j.psyneuen.2016.12.007.

Felling, R.J., Singer, H.S., 2011. Neurobiology of Tourette syndrome: Current status and need for further investigation. J. Neurosci. 31, 12387–12395. https://doi.org/10.1523/JNEUROSCI.0150-11.2011.

Feng, X., Deistung, A., Dwyer, M.G., Hagemeier, J., Polak, P., Lebenberg, J., Frouin, F., Zivadinov, R., Reichenbach, J.R., Schweser, F., 2017. An improved FSL-FIRST pipeline for subcortical gray matter segmentation to study abnormal brain anatomy using quantitative susceptibility mapping (QSM). Magn. Reson. Imaging 39, 110–122. https://doi.org/10.1016/j.mri.2017.02.002.

Foa, E.B., Huppert, J.D., Leiberg, S., Langner, R., Kichic, R., Hajcak, G., Salkovskis, P.M., 2002. The Obsessive-Compulsive Inventory: Development and validation of a short version. Psychol. Assess. 14, 485–496. https://doi.org/10.1037/1040-3590.14.4.485EB2002.

Fornito, A., Arnatkevičiūtė, A., Fulcher, B.D., 2019. Bridging the gap between connectome and transcriptome. Trends Cogn. Sci. 23: 34–50. https://doi.org/10.1016/j.tics.2018.10.005.

Fox, M.D., 2018. Mapping symptoms to brain networks with the human connectome. N. Engl. J. Med. 379, 2237–2245. https://doi.org/10.1056/NEJMra1706158.

Frahm, J., Haase, A., Matthaei, D., 1986. Rapid three-dimensional MR imaging using the FLASH technique. J. Comput. Assist. Tomogr. 10, 363–368. https://doi.org/10.1097/00004728-198603000-00046.

Ganos, C., Al-Fatly, B., Fischer, J.-F., Baldermann, J.-C., Hennen, C., Visser-Vandewalle, V., Neudorfer, C., Martino, D., Li, J., Bouwens, T., Ackermanns, L., Leentjens, A.F.G., Pyatigorskaya, N., Worbe, Y., Fox, M.D., Kühn, A.A., Horn, A., 2022. A neural network for tics: Insights from causal brain lesions and deep brain stimulation. Brain 145, 4385–4397. https://doi.org/10.1093/brain/awac009.

Gerasch, S., Kanaan, A.S., Jakubovski, E., Müller-Vahl, K. R., 2016. Aripiprazole improves associated comorbid conditions in addition to tics in adult patients with Gilles de La Tourette syndrome. Front. Neurosci. 10, 416. https://doi.org/10.3389/fnins.2016.00416.

Ghosh, D., Burkman, E., 2017. Relationship of serum ferritin level and tic severity in children with Tourette syndrome. Child’s Nerv. Syst. 33, 1373–1378. https://doi.org/10.1007/s00381-017-3424-z.

Goellner, B., Aberle, H., 2012. The synaptic cytoskeleton in development and disease. Dev. Neurobiol. 72, 111–125. https://doi.org/10.1002/dneu.20892.

Goetz, C.G., Pappert, E.J., Louis, E.D., Raman, R., Leurgans, S., 1999. Advantages of a modified scoring method for the Rush Video-Based Tic Rating Scale. Mov. Disord. 14, 502–506. https://doi.org/10.1002/1531-8257(199905)14:3<502::AID-MDS1020>3.0.CO;2-G.

Goodman, W.K., Price, L.H., Rasmussen, S.A., Mazure, C., Fleischmann, R.L., Hill, C.L., Heninger, G.R., Charney, D.S., 1989. The Yale-Brown Obsessive Compulsive Scale. I. Development, use, and reliability. Arch. Gen. Psychiatry 46, 1006–1011. https://doi.org/10.1001/archpsyc.1989.01810110048007.

Gorman, D.A., Zhu, H., Anderson, G.M., Davies, M., Peterson, B.S., 2006. Ferritin levels and their association with regional brain volumes in Tourette’s syndrome. Am. J. Psychiatry 163, 1264–1272. https://doi.org/10.1176/appi.ajp.163.7.1264.

Grace, A.A., Floresco, S.B., Goto, Y., Lodge, D.J., 2007. Regulation of firing of dopaminergic neurons and control of goal-directed behaviors. Trends Neurosci. 30, 220–227. https://doi.org/10.1016/j.tins.2007.03.003.

Guo, Y., Su, L., Zhang, J., Lei, J., Deng, X., Xu, H., Yang, Z., Kuang, S., Tang, J., Luo, Z., Deng, H., 2012. Analysis of the *BTBD9* and *HTR2C* variants in Chinese Han patients with Tourette syndrome. Psychiatr. Genet. 22, 300–303. https://doi.org/10.1097/YPG.0b013e32835862b1

Haacke, E.M., Lenz, G.W., 1987. Improving MR image quality in the presence of motion by using rephasing gradients. AJR Am. J. Roentgenol. 148, 1251–1258. https://doi.org/10.2214/ajr.148.6.1251.

Hallgren, B., Sourander, P., 1958. The effect of age on the non-haemin iron in the human brain. J. Neurochem. 3, 41–51. https://doi.org/10.1111/j.1471-4159.1958.tb12607.x.

Hawrylycz, M.J., Lein, E.S., Guillozet-Bongaarts, A.L., Shen, E.H., Ng, L., Miller, J.A., van de Lagemaat, L.N., Smith, K.A., Ebbert, A., Riley, Z.L., Abajian, C., Beckmann, C.F., Bernard, A., Bertagnolli, D., Boe, A.F., Cartagena, P.M., Chakravarty, M.M., Chapin, M., Chong, J., Dalley, R.A., Daly, B.D., Dang, C., Datta, S., Dee, N., Dolbeare, T.A., Faber, V., Feng, D., Fowler, D.R., Goldy, J., Gregor, B.W., Haradon, Z., Haynor, D.R., Hohmann, J.G., Horvath, S., Howard, R.E., Jeromin, A., Jochim, J.M., Kinnunen, M., Lau, C., Lazarz, E.T., Lee, C., Lemon, T.A., Li, L., Li, Y., Morris, J.A., Overly, C.C., Parker, P.D., Parry, S.E., Reding, M., Royall, J.J., Schulkin, J., Sequeira, P.A., Slaughterbeck, C.R., Smith, S.C., Sodt, A.J., Sunkin, S.M., Swanson, B.E., Vawter, M.P., Williams, D., Wohnoutka, P., Zielke, H.R., Geschwind, D.H., Hof, P.R., Smith, S.M., Koch, C., Grant, S.G.N., Jones, A.R., 2012. An anatomically comprehensive atlas of the adult human brain transcriptome. Nature 489. 391–399. https://doi.org/10.1038/nature11405.

Hill, J.M., 1988. The distribution of iron in the brain, in: Youdim, M.B.H. (Ed.), Brain Iron: Neurochemical and Behavioural Aspects. Taylor and Francis, London, UK, pp 1–25.

Hoekstra, P.J., Dietrich, A., Edwards, M.J., Elamin, I., Martino, D., 2013. Environmental factors in Tourette syndrome. Neurosci. Biobehav. Rev. 37, 1040–1049. http://dx.doi.org/10.1016/j.neubiorev.2012.10.010

Jenkinson, M., Beckmann, C.F., Behrens, T.E.J., Woolrich, M.W., Smith, S.M., 2012. FSL. NeuroImage 62, 782–790. https://doi.org/10.1016/j.neuroimage.2011.09.015.

Kalanithi, P.S.A., Zheng, W., Kataoka, Y., DiFiglia, M., Grantz, H., Saper, C.B., Schwartz, M.L., Leckman, J.F., Vaccarino, F.M., 2005. Altered parvalbumin-positive neuron distribution in basal ganglia of individuals with Tourette syndrome. Proc. Natl. Acad. Sci. USA 102, 13307–13312. https://doi.org/10.1073/pnas.0502624102.

Kanaan, A.S., Gerasch, S., García-García, I., Lampe, L., Pampel, A., Anwander, A., Near, J., Möller, H.E., Müller-Vahl, K., 2017. Pathological glutamatergic neurotransmission in Gilles de la Tourette syndrome. Brain 140, 218–234. https://doi.org/10.1093/brain/aww285.

Kanaan, A.S., Anwander, A., Metere, R., Schäfer, A., Schlumm, T., Near, J., Bilgic, B., Müller-Vahl, K., Möller, H.E., 2018. Iron-related gene expression associated with magnetic susceptibility reductions: Application to the pathophysiology of a movement disorder population. Proceedings of the 26th Annual Meeting of ISMRM, Paris, France, abstract 222.

Kennedy, B.C., Wallin, D.J., Tran, P.V., Georgieff, M.K., 2016. Long-term brain and behavioral consequences of early-life iron deficiency, in: Reissland, N., Kisilevsky, B.S. (Eds.). Fetal Development: Research on Brain and Behavior, Environmental Influences, and Emerging Technologies. Springer, Cham, Switzerland, pp. 295–316. https://doi.org/10.1007/978-3-319-22023-9_15.

Korkmaz, S., Goksuluk, D., Zararsiz, G., 2014. MVN: An R package for assessing multivariate normality. R J. 6, 151–162. https://doi.org/10.32614/RJ-2014-031.

Krebs, N., Langkammer, C., Goessler, W., Ropele, S., Fazekas, F., Yen, K., Scheurer, E., 2014. Assessment of trace elements in human brain using inductively coupled plasma mass spectrometry. J. Trace Elem. Med. Biol. 28, 1–7. https://doi.org/10.1016/j.jtemb.2013.09.006.

Kuleshov MV, Jones MR, Rouillard AD, Fernandez NF, Duan Q, Wang Z, Koplev, S., Jenkins, S.L., Jagodnik, K.M., Lachmann, A., McDermott, M.G., Monteiro, C.D., Gundersen, G.W., Ma’ayan, A., 2016. Enrichr: A comprehensive gene set enrichment analysis web server 2016 update. Nucleic Acids Res. 44, W90–W97. https://doi.org/10.1093/nar/gkw377.

Langkammer C, Schweser F, Krebs N, Deistung A, Goessler W, Scheurer E, Sommer, K., Reishofer, G., Yen, K., Fazekas, F., Ropele, S., Reichenbach, J.R., 2012. Quantitative susceptibility mapping (QSM) as a means to measure brain iron? A post mortem validation study. NeuroImage 62, 1593– 1599. https://doi.org/10.1016/j.neuroimage.2012.05.049.

Larsen, B., Olafsson, V., Calabro, F., Laymon, C., Tervo-Clemmens, B., Campbell, E., Minhas, D., Montez, D., Price, J., Luna, B., 2020. Maturation of the human striatal dopamine system revealed by PET and quantitative MRI. Nat. Commun. 11, 846. https://doi.org/10.1038/s41467-020-14693-3.

Leckman, J.F., 2002. Tourette’s syndrome. Lancet 360, 1577–1586. https://doi.org/10.1016/S0140-6736(02)11526-1.

Leckman, J.F., Riddle, M.A., Hardin, M.T., Ort, S.I., Swartz, K.L., Stevenson, J., Cohen, D.J., 1989. The Yale Global Tic Severity Scale: Initial testing of a clinician-rated scale of tic severity. J. Am. Acad. Child Adolesc. Psychiatry 28, 566–573. https://doi.org/10.1097/00004583-198907000-00015.

Lennington, J.B., Coppola, G., Kataoka-Sasaki, Y., Fernandez, T.V., Palejev, D., Li, Y., Huttner, A., Pletikos, M., Sestan, N., Leckman, J.F., Vaccarino, F.M., 2016. Transcriptome analysis of the human striatum in Tourette syndrome. Biol. Psychiatry 79, 372–382. https://doi.org/10.1016/j.biopsych.2014.07.018.

Lerner, A., Bagic, A., Simmons, J.M., Mari, Z., Bonne, O., Xu, B., Kazuba, D., Herscovitch, P., Carson, R.E., Murphy, D.L., Drevets, W.C., Hallett, M., 2012. Widespread abnormality of the γ-aminobutyric acid-ergic system in Tourette syndrome. Brain 135, 1926–1936. https://doi.org/10.1093/brain/aws104.

Li, W., Wu, B., Batrachenko, A., Bancroft-Wu, V., Morey, R.A., Shashi, V., Langkammer, C., De Bellis, M.D., Ropele, S., Song, A.W., Liu, C., 2014. Differential developmental trajectories of magnetic susceptibility in human brain gray and white matter over the lifespan. Hum. Brain Mapp. 35, 2698–2713. https://doi.org/10.1002/hbm.22360.

Liu, C., Li, W., Tong, K.A., Yeom, K.W., Kuzminski, S., 2015. Susceptibility-weighted imaging and quantitative susceptibility mapping in the brain. J. Magn. Reson. Imaging 42, 23–41. https://doi.org/10.1002/jmri.24768.

Lozoff, B., 2011. Early iron deficiency has brain and behavior effects consistent with dopaminergic dysfunction. J. Nutr. 141, 740S–746S. https://doi.org/10.3945/jn.110.131169.

Lozoff, B., Georgieff, M.K., 2006. Iron deficiency and brain development. Semin. Pediatr. Neurol. 13, 158–165. https://doi.org/10.1016/j.spen.2006.08.004

Lozoff, B., Beard, J., Connor, J., Felt, B., Georgieff, M., Schallert, T., 2006. Long-lasting neural and behavioral effects of iron deficiency in infancy. Nutr. Rev. 64(5 Pt 2), S34–S43. https://doi.org/10.1111/j.1753-4887.2006.tb00243.x.

Lukowski AF, Koss M, Burden MJ, Jonides J, Nelson CA, Kaciroti N, Jimenez, E., Lozoff, B., 2010. Iron deficiency in infancy and neurocognitive functioning at 19 Years: Evidence of long-term deficits in executive function and recognition memory. Nutr. Neurosci. 13, 54–70. https://doi.org/10.1179/147683010X12611460763689

Marques, J.P., Kober, T., Krueger, G., van der Zwaag, W., Van de Moortele, P.-F., Gruetter, R., 2010. MP2RAGE, a self bias-field corrected sequence for improved segmentation and T_1_ mapping at high field. NeuroImage 49, 1271–1281. https://doi.org/10.1016/j.neuroimage.2009.10.002.

Metere, R., Möller, H.E., 2017. PyMRT and DCMPI: Two new Python packages for MRI data analysis. Proceedings of the 25th Annual Meeting of ISMRM, Honolulu, HI, USA, abstract 3816.

Mink, J.W., 1996. The basal ganglia: Focused selection and inhibition of competing motor programs. Prog. Neurobiol. 50, 381–425. https://doi.org/10.1016/S0301-0082(96)00042-1.

Möller, H.E., Bossoni, L., Connor, J.R., Crichton, R.R., Does, M.D., Ward, R.J., Zecca, L., Zucca, F.A., Ronen, I., 2019. Iron, myelin, and the brain: Neuroimaging meets neurobiology. Trends Neurosci. 42, 384–401. https://doi.org/10.1016/j.tins.2019.03.009.

Montgomery, S.A., Åsberg, M., 1979. A new depression scale designed to be sensitive to change. Br. J. Psychiatry 134, 382–389. https://doi.org/10.1192/bjp.134.4.382.

Morgan, S.E., Seidlitz, J., Whitaker, K.J., Romero-Garcia, R., Clifton, N.E., Scarpazza, C., van Amelsvoort, T., Marcelis, M., van Os, J., Donohoe, G., Mothersill, D., Corvin, A., Andrew Pocklington, A., Raznahan, A., Philip McGuire, P., Vértes, P.E., Bullmore, E.T., 2019. Cortical patterning of abnormal morphometric similarity in psychosis is associated with brain expression of schizophrenia-related genes. Proc. Natl. Acad. Sci. USA 116, 9604–9609. https://doi.org/10.1073/pnas.1820754116.

Mortamet, B., Bernstein, M.A., Jack, Jr., C.R., Gunter, J.L., Ward, C., Britson, P.J., Meuli, R., Thiran, J.-P., Krueger, G., Alzheimer’s Disease Neuroimaging Initiative, 2009. Automatic quality assessment in structural brain magnetic resonance imaging. Magn. Reson. Med. 62, 365–372. https://doi.org/10.1002/mrm.21992.

Müller-Vahl, K.R., Bindila, L., Lutz, B., Musshoff, F., Skripuletz, T., Baumgaertel, C., Sühs, K.-W., 2020. Cerebrospinal fluid endocannabinoid levels in Gilles de la Tourette syndrome. Neuropsychopharmacology 45, 1323–1329. https://doi.org/10.1038/s41386-020-0671-6.

Palminteri, S., Lebreton, M., Worbe, Y., Grabli, D., Hartmann, A., Pessiglione, M., 2009. Pharmacological modulation of subliminal learning in Parkinson’s and Tourette’s syndromes. Proc. Natl. Acad. Sci. USA 106, 19179–19184. https://doi.org/10.1073/pnas.0904035106.

Patenaude, B., Smith, S.M., Kennedy, D.N., Jenkinson, M., 2011. A Bayesian model of shape and appearance for subcortical brain segmentation. NeuroImage 56, 907–922. https://doi.org/10.1016/j.neuroimage.2011.02.046.

Persson, N., Wu, J., Zhang, Q., Liu, T., Shen, J., Bao, R., Ni, M., Liu, T., Wang, Y., Spincemaille, P., 2015. Age and sex related differences in subcortical brain iron concentrations among healthy adults. NeuroImage 122, 385–398. http://doi.org/10.1016/j.neuroimage.2015.07.050.

Peterson, B.S., Gore, J.C., Riddle, M.A., Cohen, D.J., Leckman, J.F., 1994. Abnormal magnetic resonance imaging T_2_ relaxation time asymmetries in Tourette’s syndrome. Psychiatry Res. 55, 205–221. https://doi.org/10.1016/0165-1781(95)91246-A.

Rice, M.E., Patel, J.C., Cragg, S.J., 2011. Dopamine release in the basal ganglia. Neuroscience 198, 112–137. https://doi.org/10.1016/j.neuroscience.2011.08.066.

Rivière J-B, Xiong L, Levchenko A, St-Onge J, Gaspar C, Dion Y, Lespérance, P., Tellier, G., Richer, F., Chouinard, S., Rouleau, G.A., the Montreal Tourette Study Group, 2009. Association of intronic variants of the BTBD9 gene with Tourette syndrome. Arch. Neurol. 66, 1267–1272. https://doi.org/10.1001/archneurol.2009.213.

Romero-Garcia, R., Warrier, A., Bullmore, E.T., Baron-Cohen, S., Bethlehem, R.A.I., 2019. Synaptic and transcriptionally downregulated genes are associated with cortical thickness differences in autism. Mol, Psychiatry 24, 1053–1064. https://doi.org/10.1038/s41380-018-0023-7.

Sabbir, M.G., 2018. Loss of Ca^2+^/calmodulin dependent protein kinase kinase 2 leads to aberrant transferrin phosphorylation and trafficking: A potential biomarker for Alzheimer’s disease. Front. Mol. Biosci. 5, 1–28. https://doi.org/10.3389/fmolb.2018.00099.

Sabbir, M.G., 2020. CAMKK2-CAMK4 signaling regulates transferrin trafficking, turnover, and iron homeostasis. Cell Commun. Signal. 18, 1–21. https://doi.org/10.1186/s12964-020-00575-0.

Saß, H., Wittchen, H.-U., Zaudig, M., Houben, I., 2003. Diagnostisches und Statistisches Manual Psychischer Störungen – Textrevision – DSM-IV-TR. Hofgrefe, Göttingen, Germany.

Scharf, J.M., Yu, D., Mathews, C.A., Neale, B.M., Steward, S.E., Fagerness, J.A., Evans, P., Gamazon, E., Edlund, C.K., Service, S.K., Tikhomirov, A., Osiecki, L., Illmann, C., Pluzhnikov, A., Konkashbaev, A., Davis, L.K., Han, B., Crane, J., Moorjani, P., Crenshaw, A.T., Parkin, M.A., Reus, V.I., Lowe, T.L., Rangel-Lugo, M., Chouinard, S., Dion, Y., Girard, S., Cath, D.C., Smit, J.H., King, R.A., Fernandez, T.V., Leckman, J.F., Kidd, K.K., Kidd, J.R., Pakstis, A.J., State, M.W., Herrera, L.D., Romero, R., Fournier, E., Sandor, P., Barr, C.L., Phan, N., Gross-Tsur, V., Benarroch, F., Pollak, Y., Budman, C.L., Bruun, R.D., Erenberg, G., Naarden, A.L., Lee, P.C., Weiss, N., Kremeyer, B., Berrío, G.B., Campbell, D.D., Cardona Silgado, J.C., Ochoa, W.C., Mesa Restrepo, S.C., Muller, H., Valencia Duarte, A.V., Lyon, G.J., Leppert, M., Morgan, J., Weiss, R., Grados, M.A., Anderson, K., Davarya, S., Singer, H., Walkup, J., Jankovic, J., Tischfield, J.A., Heiman, G.A., Gilbert, D.L., Hoekstra, P.J., Robertson, M.M., Kurlan, R., Liu, C., Gibbs, J.R., Singleton, A., 22ort he North American Brain Expression Consortium, Hardy, F., 22ort he UK Human Brain Expression Database, Strengman, E., Ophoff, R.A., Wagner, M., Moessner, R., Mirel, D.B., Posthuma, D., Sabatti, C., Eskin, E., Conti, D.V., Knowles, J.A., Ruiz-Linares, A. Rouleau, G.A., Purcell, S., Heutink, P., Oostra, B.A., McMahon, W.M., Freimer, N.B., Cox, N.J., Pauls, D.L., 2013. Genome-wide association study of Tourette’s syndrome. Mol. Psychiatry 18, 721–728. https://doi.org/10.1038/mp.2012.69.

Schweser, F., Deistung, A., Lehr, B.W., Reichenbach, J.R., 2011. Quantitative imaging of intrinsic magnetic tissue properties using MRI signal phase: An approach to in vivo brain iron metabolism? NeuroImage 54, 2789–2807. https://doi.org/10.1016/j.neuroimage.2010.10.070.

Schweser, F., Deistung, A., Sommer, K., Reichenbach, J.R., 2013. Toward online reconstruction of quantitative susceptibility maps: Superfast dipole inversion. Magn. Reson. Med. 69, 1581–1593. https://doi.org/10.1002/mrm.24405.

Singer, H.S., 2013. The neurochemistry of Tourette syndrome, in: Martino, D., Leckman, J.F. (Eds.), Tourette Syndrome. Oxford University Press, Oxford, UK, pp. 276–297. https://doi.org/10.1093/med/9780199796267.001.0001.

Stankiewicz, J., Panter, S.S., Neema, M., Arora, A., Batt, C.E., Bakshi, R., 2007. Iron in chronic brain disorders: Imaging and neurotherapeutic implications. Neurotherapeutics 4, 371–386. https://doi.org/10.1016/j.nurt.2007.05.006.

Straub, S., Schneider, T.M., Emmerich, J., Freitag, M.T., Ziener, C.H., Schlemmer, H.-P., Ladd, M.E., Laun, F.B., 2017. Suitable reference tissues for quantitative susceptibility mapping of the brain. Magn. Reson. Med. 78, 204–214. https://doi.org/10.1002/mrm.26369.

Streitbürger, D.-P., Pampel, A., Krueger, G., Lepsien, J., Schroeter, M.L., Mueller, K., Möller, H.E., 2014. Impact of image acquisition on voxel-based-morphometry investigations of age-related structural brain changes. NeuroImage 87, 170–182. https://doi.org/10.1016/j.neuroimage.2013.10.051.

Szklarczyk, D., Gable, A.L., Lyon, D., Junge, A., Wyder, S., Huerta-Cepas, J., Simonovic, M., Doncheva, N.T., Morris, J.H., Bork, P., Jensen, L.J., von Mering C., 2019. STRING v11: Protein-protein association networks with increased coverage, supporting functional discovery in genome-wide experimental datasets. Nucleic Acids Res. 47, D607–D613. https://doi.org/10.1093/nar/gky1131.

Thomas, G.E.C., Leyland, L.A., Schrag, A.-E., Lees, A.J., Acosta-Cabronero, J., Weil, R.S., 2020. Brain iron deposition is linked with cognitive severity in Parkinson’s disease. J. Neurology Neurosurg. Psychiatry 91, 418–425. https://doi.org/10.1136/jnnp-2019-322042.

Thomas, G.E.C., Zarkali, A., Ryten, M., Shmueli, K., Gil-Martinez, A.L., Leyland, L.-A., McColgan, P., Acosta-Cabronero, J., Lees, A.J., Weil, R.S., 2021. Regional brain iron and gene expression provide insights into neurodegeneration in Parkinson’s disease. Brain 144, 1787–1798. https://doi.org/10.1093/brain/awab084.

Tinaz, S., Belluscio, B.A., Malone, P., van der Veen, J.W., Hallett, M., Horovitz, S.G., 2014. Role of the sensorimotor cortex in Tourette syndrome using multimodal imaging. Hum. Brain Mapp. 35, 5834–5846. https://doi.org/10.1002/hbm.22588.

Tremblay, L., Worbe, Y., Thobois, S., Sgambato-Faure, V., Féger, J., 2015. Selective dysfunction of basal ganglia subterritories: From movement to behavioral disorders. Mov. Disord. 30, 1155–1170. https://doi.org/10.1002/mds.26199.

Uecker, M., Lai, P., Murphy, M.J., Virtue, P., Elad, M., Pauly, J.M., Vasanawala, S.S., Lustig, M., 2014. ESPIRiT—An Eigenvalue Approach to Autocalibrating Parallel MRI: Where SENSE Meets GRAPPA. Magn. Reson. Med. 71, 990–1001. https://doi.org/10.1002/mrm.24751.

Uecker, M., Holme, C., Blumenthal, M., Wang, X., Tan, Z., Scholand, N., Iyer, S., Tamir, J., Lustig, M., 2021. Mrirecon/bart: version 0.7.00. Zenodo. https://doi.org/10.5281/zenodo.4570601.

Vértes, P.E., Rittman, T., Whitaker, K.J., Romero-Garcia, R., Váša, F., Kitzbichler, M.G., Wagstyl, K., Fonagy, P., Dolan, R.J., Jones, P.B., Goodyer, I.M., the NSPN Consortium, Bullmore, E.T., 2016. Gene transcription profiles associated with inter-modular hubs and connection distance in human functional magnetic resonance imaging networks. Phil. Trans. R. Soc. B 371, 20150362. https://doi.org/10.1098/rstb.2015.0362.

Visser, E., Keuken, M.C., Forstmann, B.U., Jenkinson, M., 2016. Automated segmentation of the substantia nigra, subthalamic nucleus and red nucleus in 7 T data at young and old age. NeuroImage 139, 324–336. https://doi.org/10.1016/j.neuroimage.2016.06.039.

Wallstein, N., Pöppl, A., Capucciati, A., Pampel, A., Jäger, C., Zucca, F.A., Monzani, E., Casella, L., Zecca, L., Möller, H.E., 2022. Interplay of iron and copper in the neuromelanin-related paramagnetic relaxation enhancement. Proceedings of the 30th Annual Meeting of ISMRM, London, UK, abstract 450.

Ward KL, Tkac I, Jing Y, Felt B, Beard J, Connor J, Schallert, T., Georgieff, M.K., Rao, R., 2007. Gestational and lactational iron deficiency alters the developing striatal metabolome and associated behaviors in young rats. J. Nutr. 137, 1043–1049. https://doi.org/10.1093/jn/137.4.1043.

Wharton, S., Schäfer, A., Bowtell, R., 2010. Susceptibility mapping in the human brain using threshold-based *k*-space division. Magn. Reson. Med. 63, 1292–1304. https://doi.org/10.1002/mrm.22334.

Whitaker, K.J., Vértes, P.E., Romero-Garcia, R., Váša, F., Moutoussis, M., Prabhu, G., Weiskopf, N., Callaghan, M.F., Konrad Wagstyl, K., Rittman, T., Tait, R., Ooi, C., Suckling, J., Inkster, B., Fonagy, P., Dolan, R.J., Jones, P.B., Goodyer, I.M., the NSPN Consortium, Bullmore. E.T., 2016. Adolescence is associated with genomically patterned consolidation of the hubs of the human brain connectome. Proc. Natl. Acad. Sci. USA 113, 9105–9110. https://doi.org/10.1073/pnas.1601745113.

Worbe, Y., Palminteri, S., Hartmann, A., Vidailhet, M., Lehéricy, S., Pessiglione, M., 2011. Reinforcement learning and Gilles de la Tourette syndrome. Dissociation of clinical phenotype and pharmacological treatments. Arch. Gen. Psychiatry 68, 1257–1266. https://doi.org/10.1001/archgenpsychiatry.2011.137.

Woods, D.W., Piacentini, J., Himle, M.B., Chang, S., 2005. Premonitory urge for tics scale (PUTS): Initial psychometric results and examination of the premonitory urge phenomenon in youths with tic disorders. J. Dev. Behav. Pediatr. 26, 397–403. https://doi.org/10.1097/00004703-200512000-00001.

Yehuda, S., 1990. Neurochemical basis of behavioral effects of brain iron deficiency in animals, in: Dobbing, J. (Ed.), Brain, Behavior, and Iron in the Infant Diet. Springer, London, UK, pp. 63–76. https://doi.org/10.1007/978-1-4471-1766-7_7.

Yehuda, S., Mostofsky, D.I., 2010. Iron Deficiency and Overload: From Basic Biology to Clinical Medicine. Humana Press, New York, NY. https://doi.org/10.1007/978-1-59745-462-9.

Youdim, M.B.H., Ben-Shachar, D., Yehuda, S., 1989. Putative biological mechanisms of the effect of iron deficiency on brain biochemistry and behavior. Am. J. Clin. Nutr. 50, 607–617. https://doi.org/10.1093/ajcn/50.3.607.

Yu, D., Sul, J.H., Tsetsos, F., Nawaz, M.S., Huang, A.Y., Zelaya, I., Cornelia Illmann, C., Osiecki, L., Darrow, S.M., Hirschtritt, M.E., Greenberg, E., Muller-Vahl, K.R., Stuhrmann, M., Yves Dion, Y., Rouleau, G., Aschauer, H., Stamenkovic, M., Schlögelhofer, M., Sandor, P., Barr, C.L., Grados, M., Singer, H.S., Nöthen, M.M., Hebebrand, J., Hinney, A., King, R.A., Fernandez, T.V., Barta, C., Tarnok, Z., Nagy, P., Depienne, C., Worbe, Y., Andreas Hartmann, A., Budman, C.L., Rizzo, R., Lyon, G.J., McMahon, W.M., Batterson, J.R., Cath, D.C., Malaty, I.A., Okun, M.S., Berlin, C., Woods, D.W., Lee, P.C., Jankovic, J., Robertson, M.M., Gilbert, D.L., Brown, L.W., Coffey, B.J., Dietrich, A., Hoekstra, P.J., Kuperman, S., Zinner, S.H., Luðvigsson, P., Sæmundsen, E., Thorarensen, Ó., Atzmon, G., Barzilai, N., Wagner, M., Moessner, R., Ophoff, R., Pato, C.N., Pato, M.T., Knowles, J.A., Roffman, J.L., Smoller, J.W., Buckner, R.L., Willsey, A.J., Tischfield, J.A., Heiman, G.A., Stefansson, H., Stefansson, K., Posthuma, D., Cox, N.J., Pauls, D.L., Freimer, N.B., Neale, B.M., Davis, L.K., Paschou, P., Coppola, G., Mathews, C.A., Scharf, J.M., on behalf of the Tourette Association of America International Consortium for Genetics, the Gilles de la Tourette GWAS Replication Initiative, the Tourette International Collaborative Genetics Study, the Psychiatric Genomics Consortium Tourette Syndrome Working Group, 2019. Interrogating the genetic determinants of Tourette’s syndrome and other tic disorders through genome-wide association studies. Am. J. Psychiatry 176, 217–227. https://doi.org/10.1176/appi.ajp.2018.18070857.

Zhang Y, Chen K, Sloan SA, Bennett ML, Scholze AR, O’Keeffe S, Phatnani, H.P., Guarnieri, P., Caneda, C., Ruderisch, N., Deng, S., Liddelow, S.A., Zhang, C., Daneman, R., Maniatis, T., Barres B.A., Wu, J.Q., 2014. An RNA-sequencing transcriptome and splicing database of glia, neurons, and vascular cells of the cerebral cortex. J. Neurosci. 34, 11929–11947. https://doi.org/10.1523/JNEUROSCI.1860-14.2014. Correction: J. Neurosci. 2015; 35, 864–866. https://doi.org/10.1523/JNEUROSCI.4506-14.2015.

