## Supplementary Tables and Figures for "Convergent imaging-transcriptomic evidence for disturbed iron homeostasis in Gilles de la Tourette syndrome"

#### Table of contents

##### Abbreviations and symbols

##### Supplementary tables

Table S1 Group comparison of FLASH magnitude image quality metrics
Table S2 Group comparison of $\Delta\chi$ measurements
Table S3 Statistics of PLS regression of gene expression and case-control $\Delta\chi$ differences
Table S4 STRING PPI network statistics of downregulated and upregulated PLS1 genes

##### Supplementary figures

Figure S1 Multivariate outlier detection
Figure S2 Gene-weight distribution for PLS1 and correlation with case-control $\Delta\chi$ differences in the motor striatum
Figure S3 Gene-weight distribution for PLS1 and correlation with case-control $\Delta\chi$ differences in the executive striatum

| | |
|---|---|
| Figure S4 | Gene-weight distribution for PLS1 and correlation with case-control $\Delta\chi$ differences in the limbic striatum |
| Figure S5 | Significantly enriched GO-BP terms (ToppGene) for PLS$^-$ genes in the motor striatum against a background of brain-expressed genes |
| Figure S6 | Significantly enriched GO-CC terms (ToppGene) for PLS$^-$ genes in the motor striatum against a background of brain-expressed genes |
| Figure S7 | Significantly enriched GO-BP terms (ToppGene) for PLS$^+$ genes in the executive striatum against a background of brain-expressed genes |
| Figure S8 | Significantly enriched GO-CC terms (ToppGene) for PLS$^+$ genes in the executive striatum against a background of brain-expressed genes |
| Figure S9 | Significantly enriched GO-BP terms (ToppGene) for PLS$^+$ genes in the limbic striatum against a background of brain-expressed genes |
| Figure S10 | Significantly enriched GO-CC terms (ToppGene) for PLS$^+$ genes in the limbic striatum against a background of brain-expressed genes |

### ABBREVIATIONS AND SYMBOLS

3D = three-dimensional; ADHD = attention deficit/hyper activity disorder; AHBA = Allen human brain atlas; ANTs = Advanced Normalization Tools; BAI = Beck Anxiety Inventory; BART = Berkeley Advanced Reconstruction Toolbox; BDI-II = Beck Depression Inventory II; BP = biological process; CAARS = Conners' Adult ADHD Rating Scale; CC = cellular component; CI = confidence interval; CNR = contrast-to-noise ratio; CSF = cerebrospinal fluid; DN = dentate nucleus; DSM4 = Diagnostic and Statistical Manual of Mental Disorders, Fourth Edition; $d$ = effect size (Cohen's $d$); EFC = entropy focus criterion; ESPIRiT = Eigenvector-based Self-consistent Parallel Imaging Reconstruction—iTerative; FBR = foreground-to-background ratio; FDR = false discovery rate; FIRST = FMRIB's Integrated Registration and Segmentation Tool; FLASH = Fast Low-Angle SHot; FSL = FMRIB Software Library; FWHM = full width at half maximum; GABA = $\gamma$-aminobutyric acid; GM = gray matter; GO = gene ontology; GP = globus pallidum; GTS = Gilles de la Tourette syndrome; KEGG = Kyoto Encyclopedia of Genes and Genomes; MADRS = Montgomery Åsberg Depression Rating Scale; MF = molecular function; MNI = Montreal Neurological Institute; MP2RAGE = Magnetization-Prepared 2 RApid Gradient Echoes; MRI = magnetic resonance imaging; MS = motor tic score; $N$ = number of cases; $N_{edges}$ = number of edges; $N_{nodes}$ = number of nodes; NMDA = *N*-methyl-D-aspartate; OCB = obsessive-compulsive behavior; OCD = obsessive-compulsive disorder; OCI-R = Revised Obsessive Compulsive Inventory; OR = Fisher exact test odds ratio; $P$ = error probability; $P_{FDR}$ = FDR-adjusted error probability; PC = principal component; PC2 = second principal component; PCA = principal component analysis; PLS = partial least squares; PLS1 = first partial least squares component; $PLS^+$ = upregulated gene set; $PLS^-$ = downregulated gene set; PPI = protein-protein interaction; PUTS = Premonitory Urge for Tics Scale; PyMRT = Python Magnetic Resonance Tools; QI1 = quality index 1; QOL = quality of life scale; QSM = quantitative susceptibility mapping; RN = red nucleus; ROI = region of interest; RVTRS = Rush Video-based Tic Rating Scale; $r$ = Pearson correlation coefficient; SHARP = Sophisticated Harmonic Artifact Reduction for Phase data; SN = substantia nigra; SNR = signal-to-noise ratio; STN = subthalamic nucleus; STRING = Search Tool for Retrieval of Interacting Genes/Proteins; SVD = singular value decomposition; $T_1$ = longitudinal relaxation time; $TE$ = echo time; TKD = threshold k-space division; ToppFun = ToppGene functional annotation; $TR$ = repetition time; $t$ = $t$-value; $U$ = unbiased estimator; VS = vocal tic score; Y-BOCS = Yale-Brown Obsessive Compulsive Scale; YGTSS = Yale Global Tic Severity Scale; $Z$ = standard score; $\alpha$ = type-I error; $\Delta\chi$ = magnetic susceptibility; $\chi^2$ = $\chi^2$ distribution.

#### SUPPLEMENTARY TABLES

**Supplementary Table S1—Group comparison of FLASH magnitude image quality metrics.** Statistical tests performed on the quality metrics were inspected using FDR multiple comparison correction with significance level set at $P_{FDR} < 0.05$.

| Parameter | Controls | Patients | Cohen's *d* | CI (95%) | Statistic | *P* | $P_{FDR}$ |
|---|---|---|---|---|---|---|---|
| SNR | 2.80±0.21 | 2.72±0.24 | 0.37 | –0.05 to 0.22 | $U_{47}$=241 | 0.12 | 0.21 |
| CNR | 0.14±0.04 | 0.14±0.03 | 0.13 | –0.02 to 0.028 | $U_{47}$=276 | 0.33 | 0.4 |
| FBR | 8.19±3.22 | 6.75±2.32 | 0.5 | –0.22 to 3.12 | $U_{47}$=222 | 0.06 | 0.2 |
| EFC | 0.61±0.03 | 0.63±0.03 | –0.45 | –0.03 to 0.004 | $U_{47}$=228 | 0.08 | 0.2 |
| FWHM | 3.64±0.25 | 3.67±0.36 | –0.09 | –0.21 to 0.15 | $U_{47}$=286 | 0.4 | 0.4 |

**Abbreviations & symbols:** CI = confidence interval; CNR = contrast-to-noise ratio; *d* = effect size (Cohen's *d*); EFC = entropy focus criterion; FBR = foreground-to-background ratio; FDR = false discovery rate; FWHM = full width at half maximum; *P* = error probability; SNR = signal-to-noise ratio; *U* = unbiased estimator.

**Supplementary Table S2—Group comparison of $\Delta\chi$ measurements.** Statistical tests performed on distinct ROIs were inspected using FDR multiple comparison correction. The significance level was set at $P_{FDR} < 0.05$.

| Region | $\Delta\chi_{ctl}$ | $\Delta\chi_{GTS}$ | Cohen's *D* | CI (95%) | Statistic | *P* | $P_{FDR}$ |
|---|---|---|---|---|---|---|---|
| RN | 116±47 | 98±35 | 0.44 | –5.9 to 42.8 | $U_{47}$=232 | 0.091 | 0.091 |
| SN | 124±25 | 110±31 | 0.49 | –2.5 to 30.4 | $U_{47}$=206 | 0.032 | **0.048** |
| STN | 69±33 | 42±31 | 0.83 | 8.3 to 46.2 | $U_{47}$=159 | 0.0026 | **0.016** |
| Striatum | 18±16 | 6±13 | 0.75 | 2.5 to 19.7 | $U_{47}$=180 | 0.0088 | **0.026** |
| GP | 88±19 | 75±20 | 0.62 | 0.9 to 23.9 | $U_{47}$=197 | 0.021 | **0.042** |
| DN | 72±31 | 52±40 | 0.57 | –0.1 to 41.4 | $U_{47}$=191 | 0.016 | **0.031** |

**Abbreviations & symbols:** CI = confidence interval; DN = dentate nucleus; *d* = effect size (Cohen's *d*); FDR = false discovery rate; GP = globus pallidus; GTS = Gilles de la Tourette syndrome; *P* = error probability; RN = red nucleus; ROI = region of interest; SD = standard deviation; SN = substantia nigra; STN = subthalamic nucleus; *U* = unbiased estimator; $\Delta\chi_{ctl}$ = magnetic susceptibility (mean ± SD) in the control cohort; $\Delta\chi_{GTS}$ = magnetic susceptibility (mean ± SD) in the patient cohort.

**Supplementary Table S3—Statistics of PLS regression of gene expression and case-control $\Delta\chi$ differences.**

| Region | Component | Explained variance | *r* | *P* |
|---|---|---|---|---|
| Motor striatum | PLS1 | 48.2% | 0.7 | $4.0\times10^{-8}$ |
| Executive striatum | PLS1 | 52.2% | 0.72 | $9.2\times10^{-9}$ |
| Limbic striatum | PLS1 | 49.1% | 0.7 | $4.7\times10^{-7}$ |

**Abbreviations & symbols:** *P* = error probability; PLS = partial least squares; PLS1 = first PLS component; *r* = Pearson correlation coefficient.

**Supplementary Table S4—STRING PPI network statistics of downregulated and upregulated PLS1 genes.**

| $PLS^{\pm}$ | $N_{nodes}$ | $N_{edges}$ | $N_{edges}$, expected | Local clustering coef.[a] | Average node degree | PPI enrichment, *P*-value |
|---|---|---|---|---|---|---|
| Motor $PLS^{-}$ | 927 | 1 046 | 793 | 0.28 | 2.26 | $<1.0\times10^{-16}$ |
| Motor $PLS^{+}$ | 928 | 1 665 | 1 314 | 0.33 | 3.59 | $<1.0\times10^{-16}$ |
| Exec. $PLS^{-}$ | 926 | 779 | 757 | 0.26 | 1.68 | 0.22 |
| Exec. $PLS^{+}$ | 904 | 1 509 | 1 278 | 0.34 | 3.34 | $1.7\times10^{-10}$ |
| Limbic $PLS^{-}$ | 950 | 901 | 733 | 0.27 | 1.9 | $1.2\times10^{-9}$ |
| Limbic $PLS^{+}$ | 909 | 745 | 593 | 0.29 | 1.64 | $1.1\times10^{-9}$ |

[a] Mean values.

**Abbreviations & symbols:** $N_{edges}$ = number of edges; $N_{nodes}$ = number of nodes; *P* = error probability; $PLS^{+}$ upregulated gene set; $PLS^{-}$ downregulated gene set; PPI = protein-protein interaction; STRING = Search Tool for Retrieval of Interacting Genes/Proteins.

#### SUPPLEMENTARY FIGURES

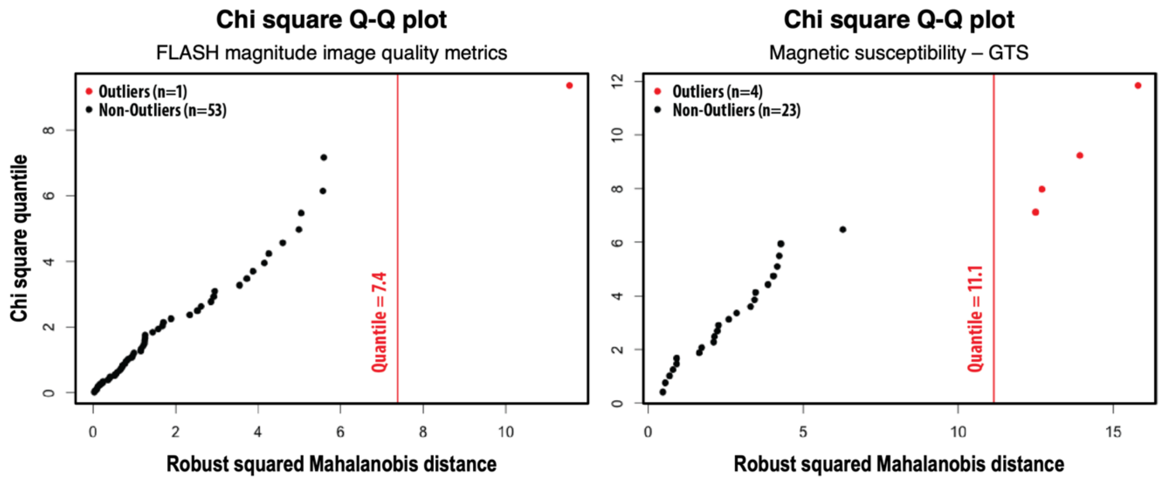


**Supplementary Figure S1—Multivariate outlier detection.** A step-wise multivariate outlier detection approach implementing a robust Mahalanobis distance framework was used to remove low-quality data that may be affected by motion. In the first step, outliers from the entire sample were detected based on structural image quality indices calculated on the magnitude image. In the second step, outliers were detected based on $\Delta\chi$ values extracted from subcortical nuclei for each sample separately. Data were regarded as outliers if the robust Mahalanobis distance was greater than the 97.5% quantile of the $\chi^2$ distribution.

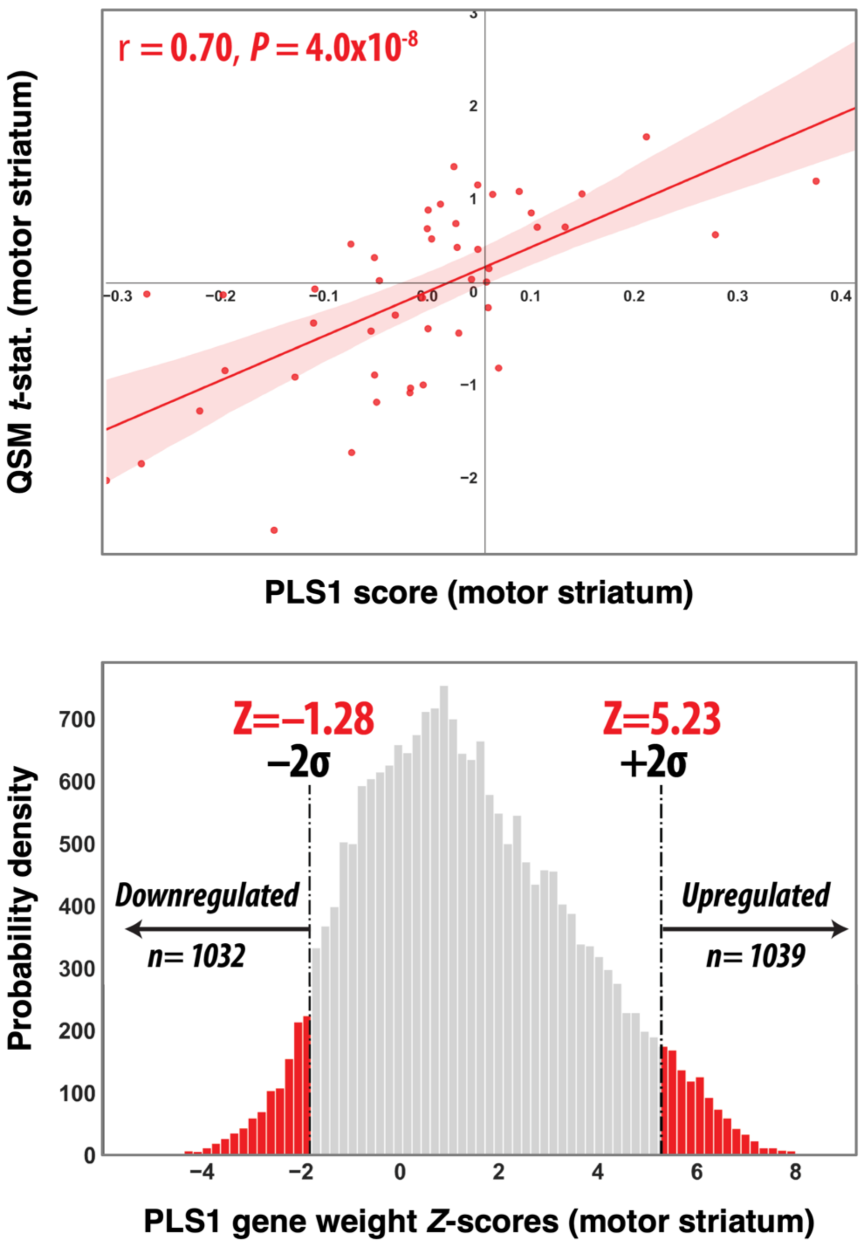


**Supplementary Figure S2—Gene-weight distribution for PLS1 and correlation with case-control $\Delta\chi$ differences in the motor striatum.** Scatterplot of regional PLS1 scores (weighted sum of 20,737 gene expression scores) vs. case-control $\Delta\chi$ differences in the motor striatum **(top)** and bootstrapped distribution of PLS1 gene *Z*-scores outlining the cutoffs for downregulated ($PLS^{-}$) and upregulated ($PLS^{+}$) genes used for PPI network and enrichment analysis **(bottom)**.

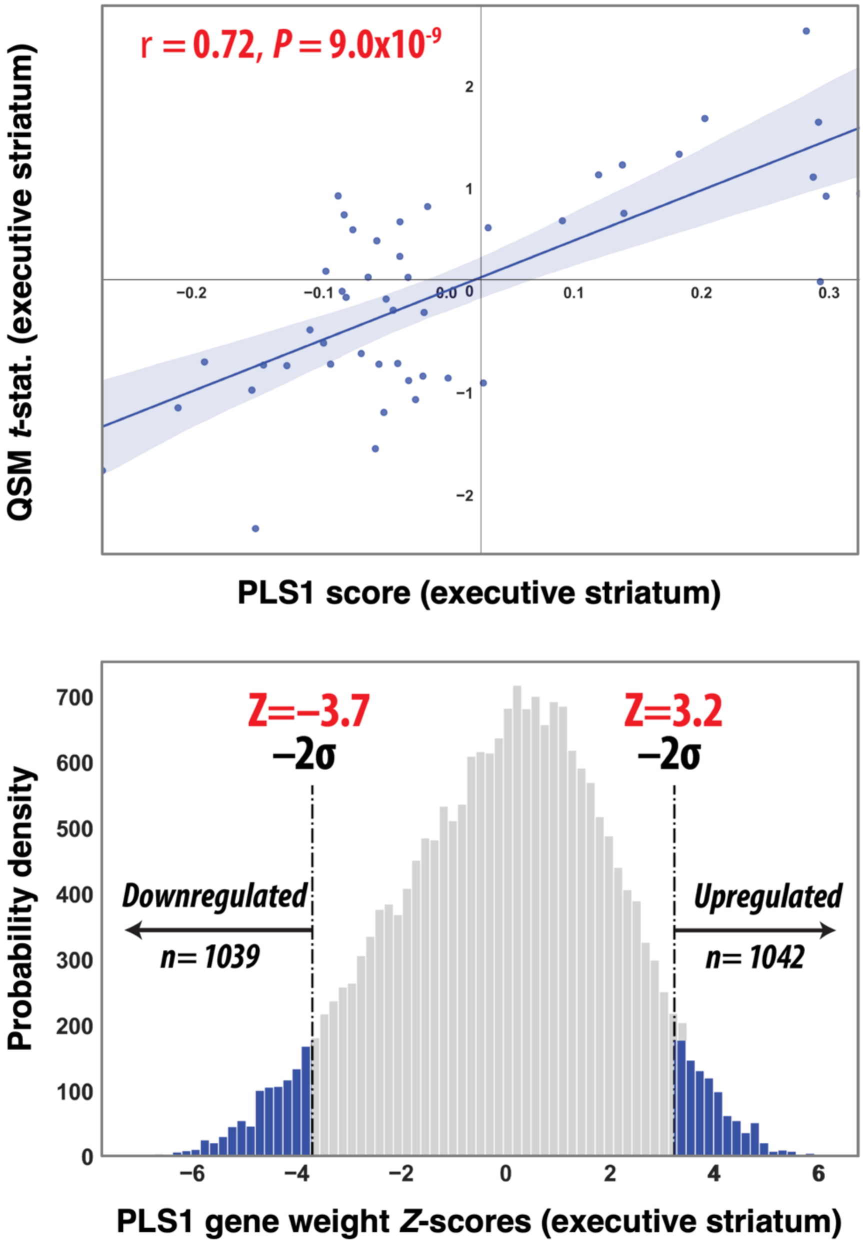


**Supplementary Figure S3—Gene-weight distribution for PLS1 and correlation with case-control $\Delta\chi$ differences in the executive striatum.** Scatterplot of regional PLS1 scores (weighted sum of 20,737 gene expression scores) vs. case-control $\Delta\chi$ differences in the executive striatum **(top)** and bootstrapped distribution of PLS1 gene *Z*-scores outlining the cutoffs for downregulated (PLS$^{-}$) and upregulated (PLS$^{+}$) genes used for PPI network and enrichment analysis **(bottom)**.

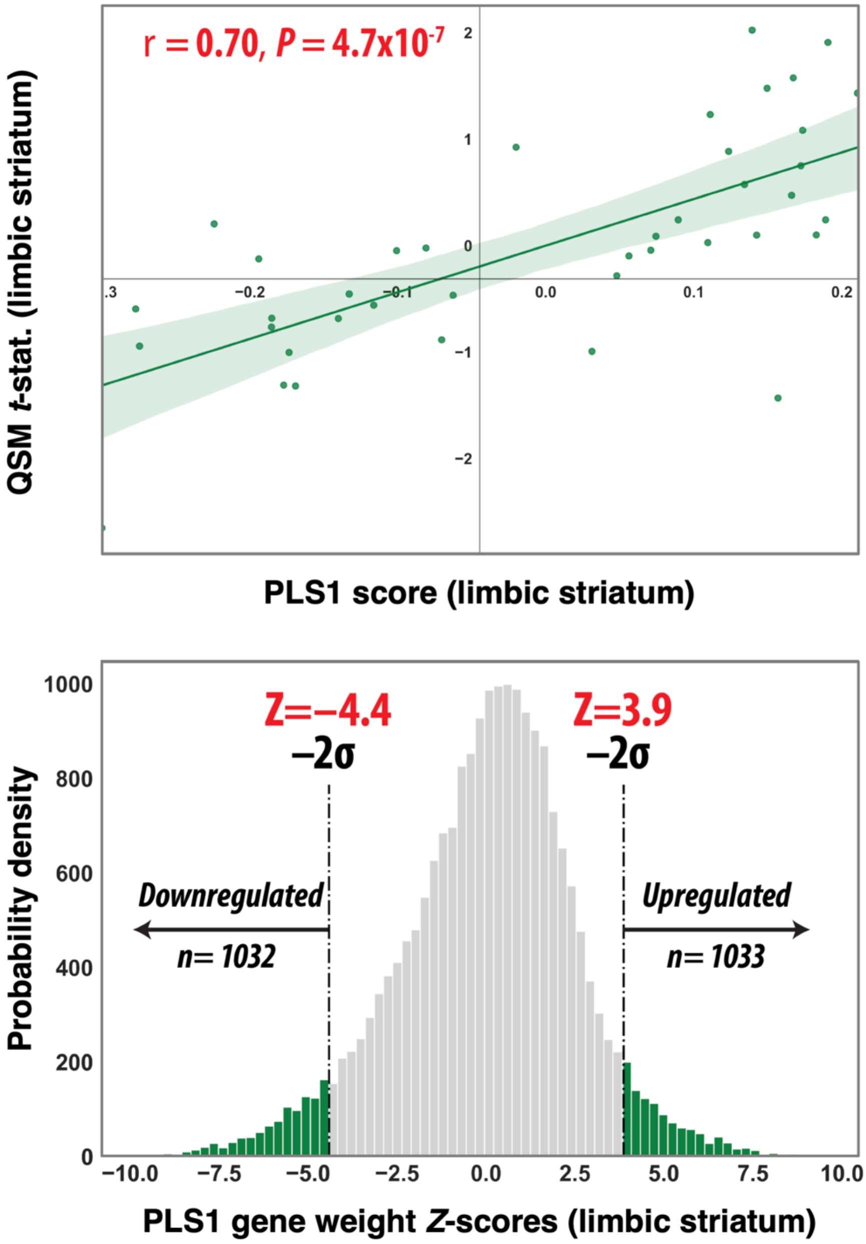


**Supplementary Figure S4—Gene-weight distribution for PLS1 and correlation with case-control Δχ differences in the limbic striatum.** Scatterplot of regional PLS1 scores (weighted sum of 20,737 gene expression scores) vs. case-control Δχ differences in the limbic striatum **(top)** and bootstrapped distribution of PLS1 gene *Z*-scores outlining the cutoffs for downregulated ($PLS^{-}$) and upregulated ($PLS^{+}$) genes used for PPI network and enrichment analysis **(bottom)**.

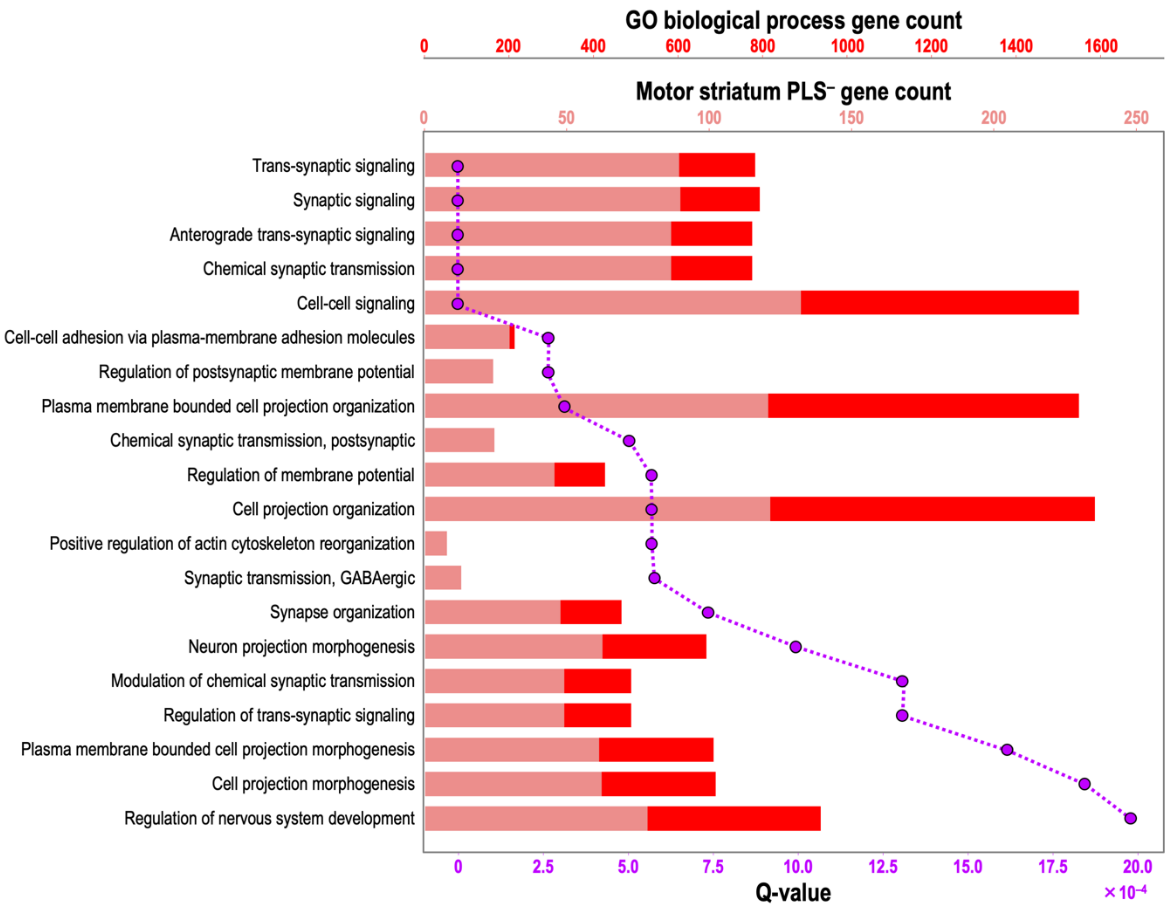


**Supplementary Figure S5—Significantly enriched GO-BP terms (ToppGene) for PLS⁻ genes in the motor striatum against a background of brain-expressed genes.** Non-overlapping enrichment terms include *“chemical synaptic transmission”* (GO-BP:0007268), *“GABAergic synaptic transmission”* (GO-BP:0051932) or *“positive regulation of actin cytoskeleton reorganization”* (GO-BP:2000251).

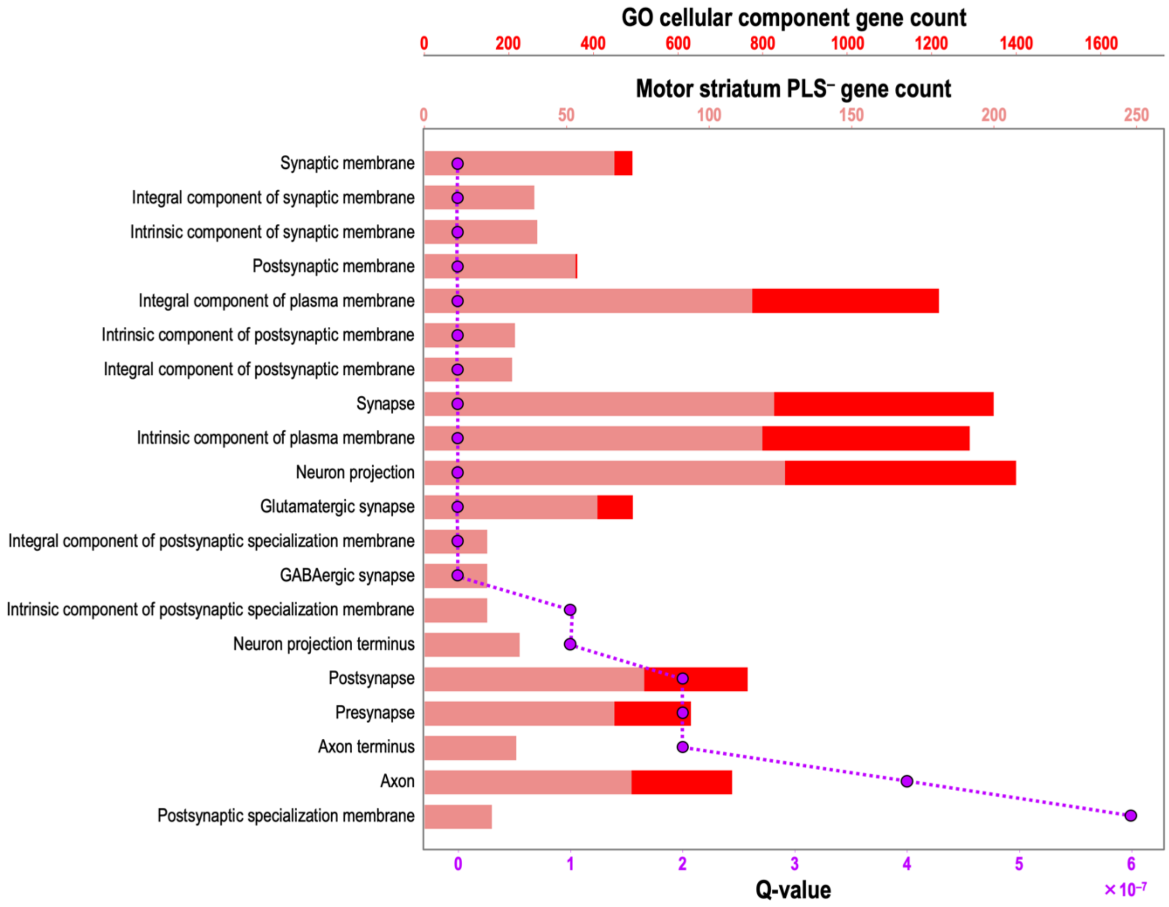


**Supplementary Figure S6—Significantly enriched GO-CC terms (ToppGene) for PLS⁻ genes in the motor striatum against a background of brain-expressed genes.** Non-overlapping enrichment terms include *"glutamatergic synapse"* (GO-CC:0098978) or *"GABA-ergic synapse"* (GO-CC:0098982).

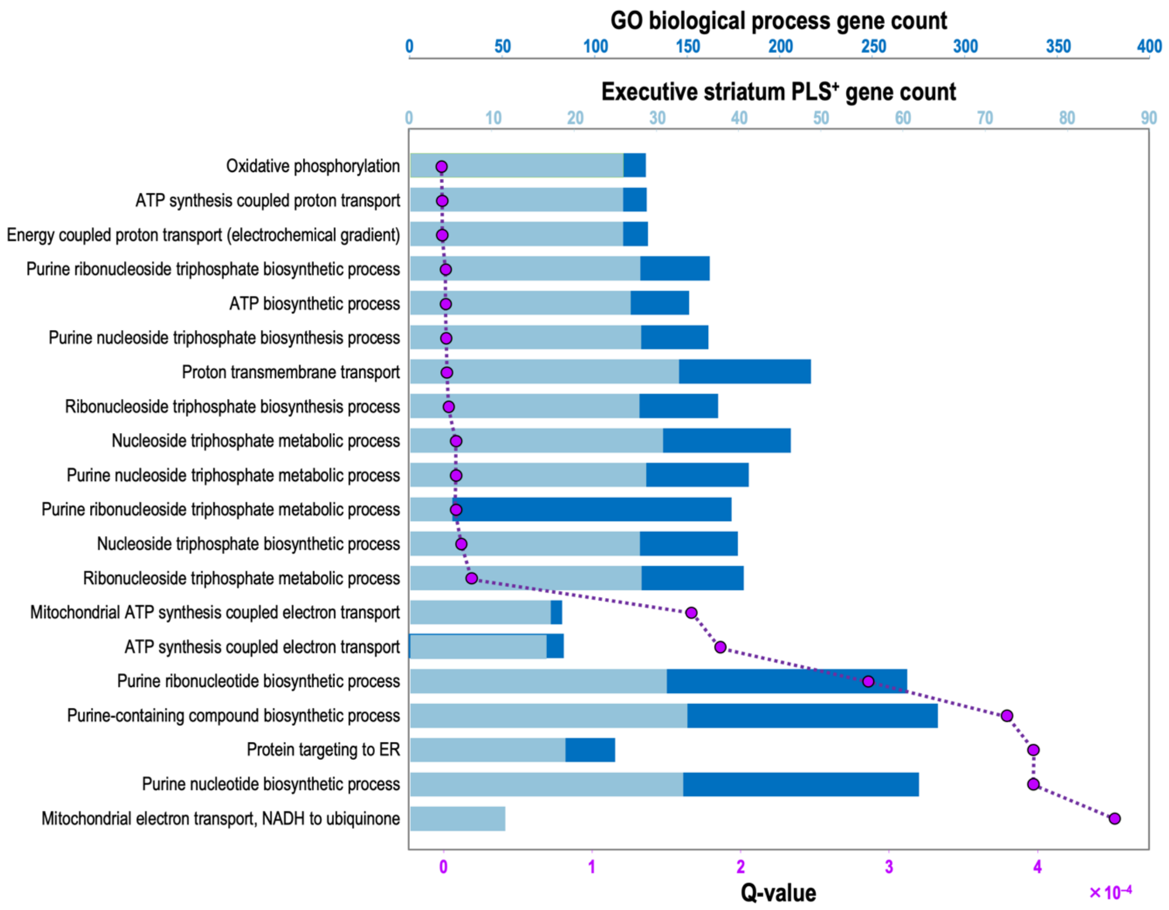


**Supplementary Figure S7—Significantly enriched GO-BP terms (ToppGene) for PLS+ genes in the executive striatum against a background of brain-expressed genes.** Enriched terms include *"mitochondrial electron transport, NADH to ubiquinone"* (GO-BP:0006120), *"oxidative phosphorylation"* (GO-BP:0006119), and *"ribonucleoside triphosphate metabolic process"* (GO-BP:0009199).

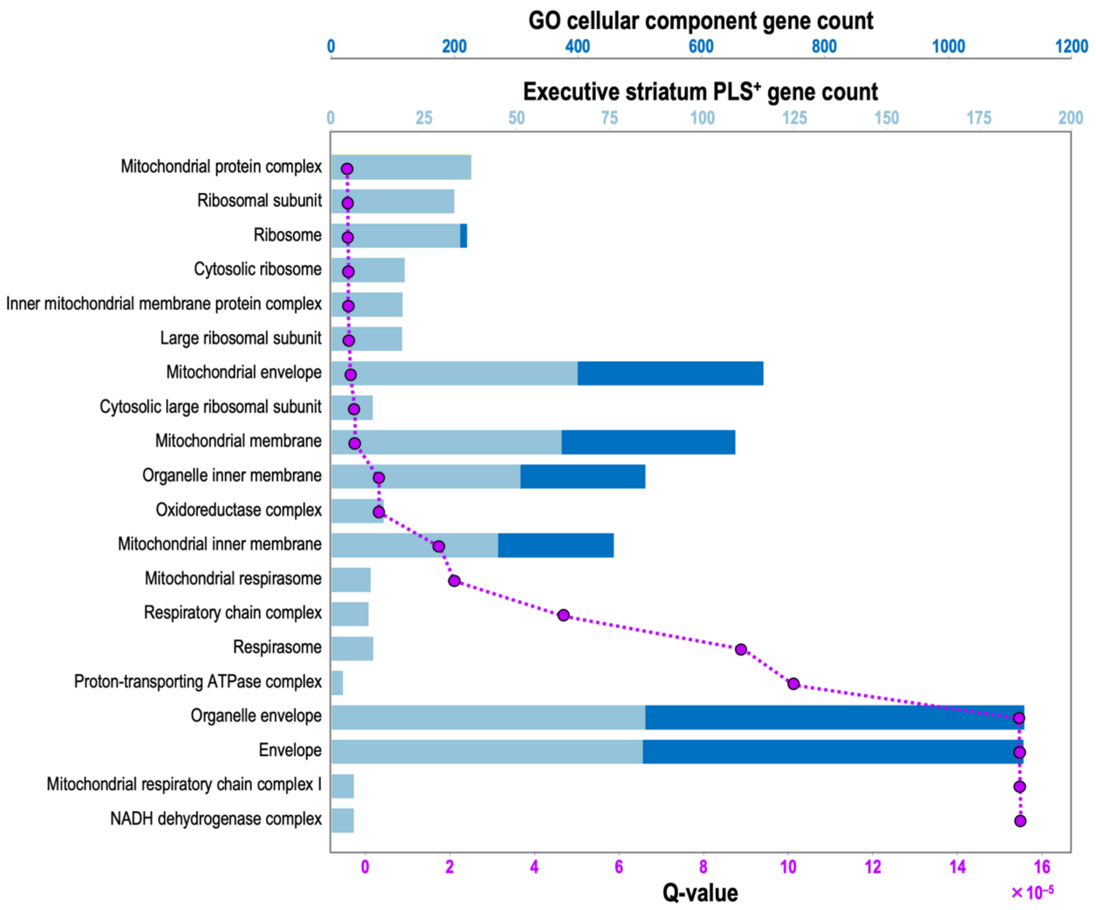


**Supplementary Figure S8—Significantly enriched GO-CC terms (ToppGene) for PLS⁺ genes in the executive striatum against a background of brain-expressed genes.** Enriched terms include *"mitochondrial protein-containing complex"* (GO-CC:0098798), *"inner mitochondrial membrane protein complex"* (GO-CC:0098800) or *"ribosomal subunit"* (GO-CC:0044391).

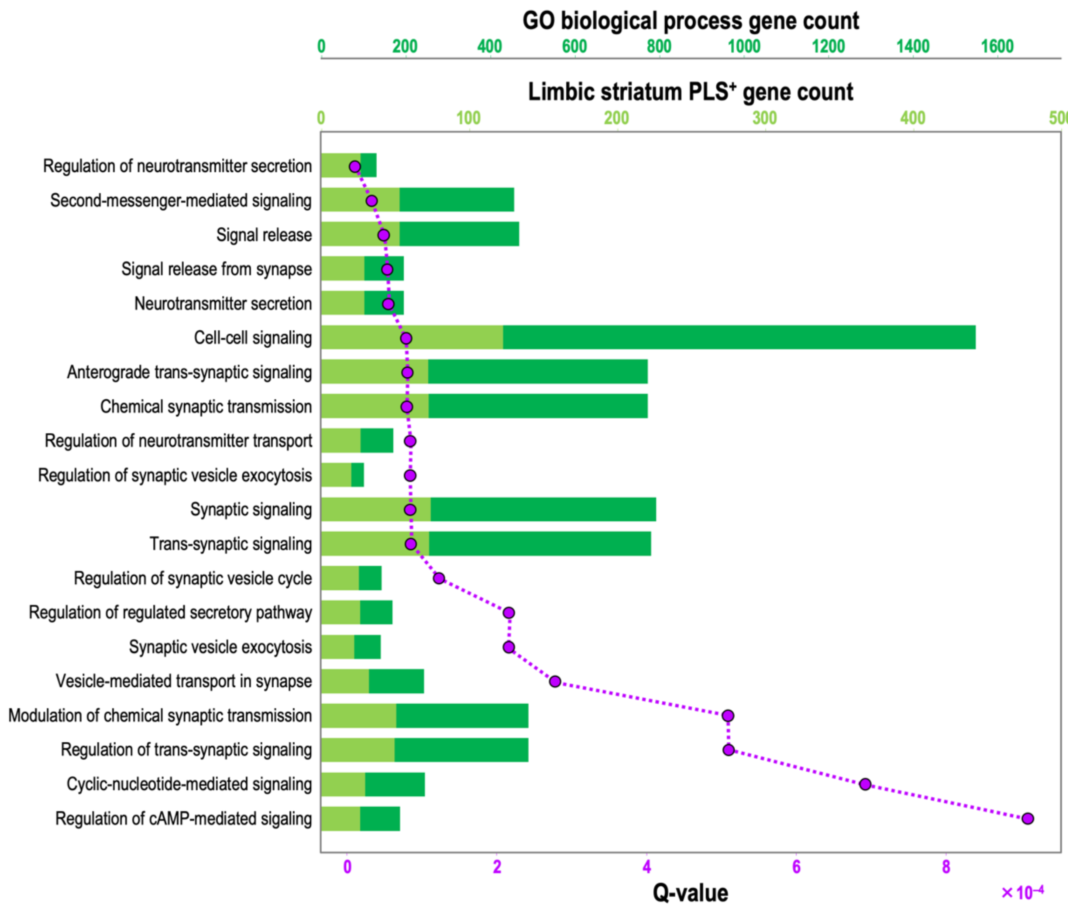


**Supplementary Figure S9—Significantly enriched GO-BP terms (ToppGene) for PLS$^+$ genes in the limbic striatum against a background of brain expressed-genes.** Enriched terms include *"second-messenger-mediated signaling"* (GO-BP:0019932) and *"regulation of neurotransmitter secretion"* (GO-BP:0046928).

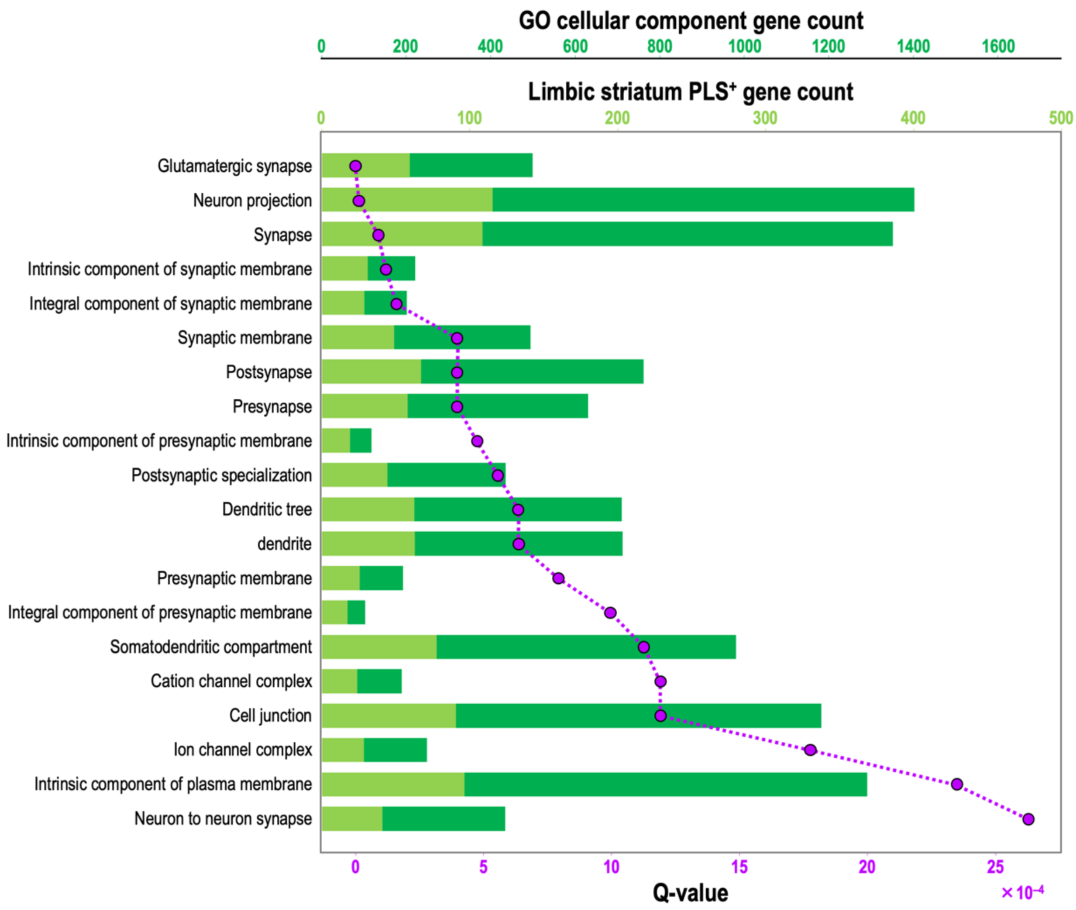


**Supplementary Figure S10—Significantly enriched GO-CC terms (ToppGene) for PLS$^{+}$ genes in the limbic striatum against a background of brain-expressed genes.** Enriched terms include *"intrinsic"* and *"integral"* synaptic membrane components.
